# Cost-effectiveness of routine COVID-19 adult vaccination programmes in England

**DOI:** 10.1101/2024.11.08.24316972

**Authors:** Matt J. Keeling, Edward M. Hill, Stavros Petrou, Phuong Bich Tran, May Ee Png, Sophie Staniszewska, Corinna Clark, Katie Hassel, Julia Stowe, Nick Andrews

## Abstract

In England, and many other countries, immunity to SARS-CoV-2 infection and COVID-19 disease is highly heterogeneous. Immunity has been acquired through natural infection, primary and booster vaccination, while protection has been lost through waning immunity and viral mutation. During the height of the pandemic in England, the main aim was to rapidly protect the population and large supplies of vaccine were pre-purchased, eliminating the need for cost-effective calculations. As we move to an era where for the majority of the population SARS-CoV-2 infections cause relatively mild disease, and vaccine stocks need to be re-purchased, it is important we consider the cost-effectiveness and economic value of COVID-19 vaccination programmes. Here using data from 2023 and 2024 in England on COVID-19 hospital admissions, ICU admissions and deaths, coupled with bespoke health economic costs, we consider the willingness to pay threshold for COVID-19 vaccines in different age and risk groups.

Willingness to pay thresholds vary from less than £1 for younger age-groups with- out any risk factors, to over £100 for older age-groups with comorbidities that place them at risk. This extreme non-linear dependence on age, means that despite the different method of estimating vaccine effectiveness, there is considerable qualitative agreement on the willingness to pay threshold, and therefore which ages it is cost-effective to vaccinate.

The historic offer of COVID-19 vaccination to those 65 and over for the autumn 2023 programme and those over 75 for the spring 2023 programme, aligns with our cost- effective threshold for pre-purchased vaccine when the only cost was administration. However, for future programmes, when vaccine costs are included, the age-thresholds slowly increase thereby demonstrating the continued importance of protecting the eldest and most vulnerable in the population.

**Graphical Abstract:** 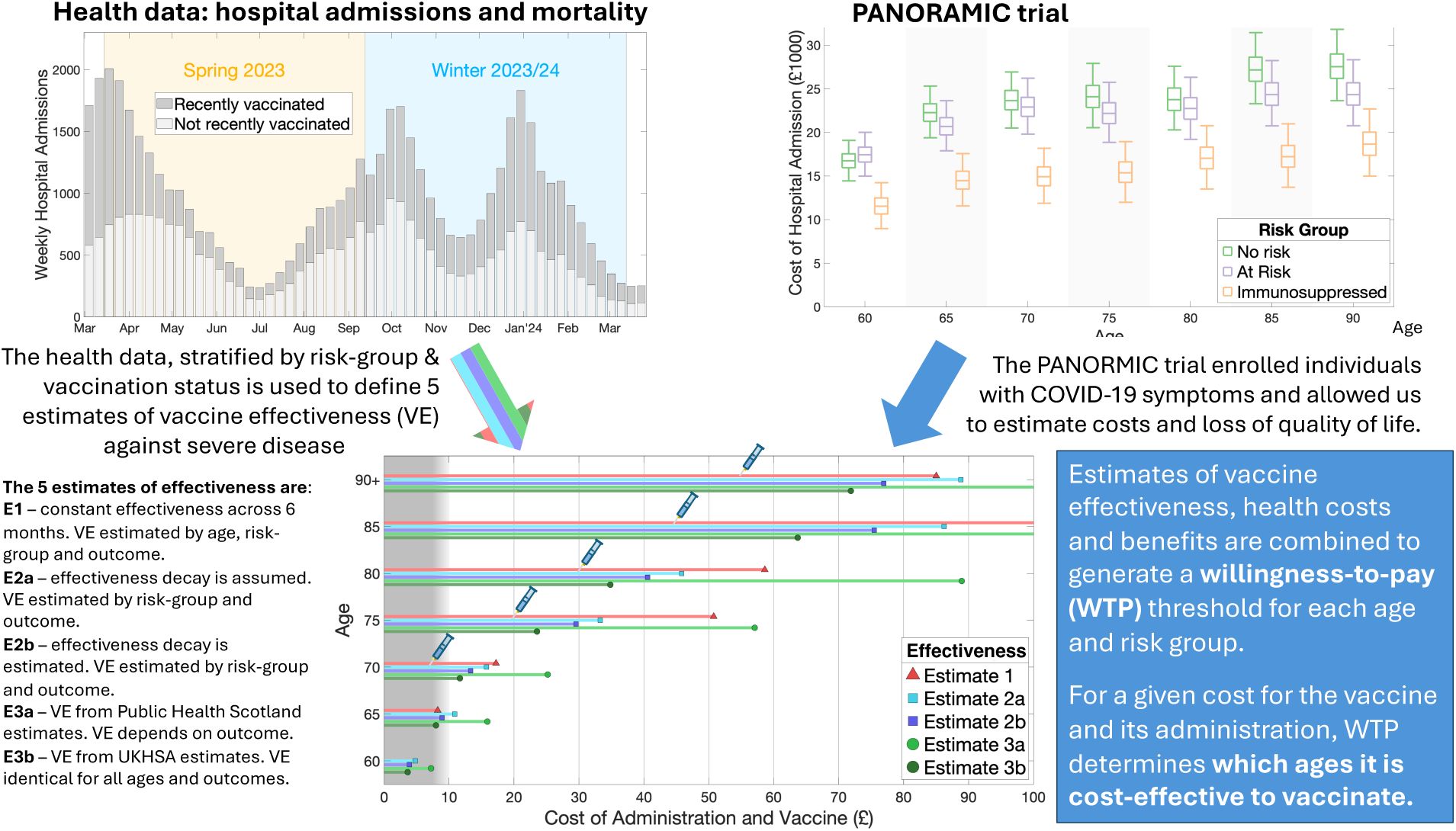

**Research In Context:** *Evidence before this study:* We searched PubMed, Google Scholar, and medRxiv for articles published in English from inception to Nov 19, 2024, with the following search terms: “COVID-19” AND “vaccin*” AND “cost-effective*”. A total of 1287 articles were identified, of which 47 considered the cost-effectiveness of vaccination against COVID-19; of these only 12 considered booster vaccination against Omicron, and only 6 considered regular vaccination. These studies focused on USA (× 2), South Korea, Thailand, Germany and the UK.

*Added value of this study:* This study provides the first independent assessment of the cost-effectiveness of the spring and autumn COVID-19 vaccination programme in the UK, and the first to consider vaccination across the spectrum of ages. It uses UK data from the Spring and Autumn boosters 2023 on severe health outcomes (hospital admission and deaths) partitioned by age, risk group and vaccine status to estimate the impact of immunisation programmes. This is combined with health economic estimates to determine the willingness-to-pay for the costs of immunisation (vaccine plus administration costs) in each age and risk group.

*Implications of all the available evidence:* Which ages and risk groups it is cost-effective to vaccinate depends on the price paid for the vaccine, although the increase in risk of severe illness with age dominates the results. Vaccination of individuals under 65 years old without additional risk factors is unlikely to be cost-effective at any vaccine price. However, universal vaccination (in both Spring and Autumn for all risk groups) for older adults is likely to be cost-effective if the vaccine can be secured for a sufficiently low price.

## 1. Introduction

The rapid development of COVID-19 specific vaccines was a major scientific triumph, which had profound implications for the control of a pandemic when population-level immunity to SARS-CoV-2 (the causative pathogen of COVID-19) was relatively low. As we move into a scenario in England where the majority of the population have been infected at least once by SARS-CoV-2 [1], and most people have had multiple primary and booster vaccine doses [2,3], it becomes important to rigorously quantify the impact and cost-effectiveness of future COVID-19 vaccination programmes. Compared to modelling and analysis of COVID-19 vaccination for England in the initial years of the pandemic [4–6], this later problem is confounded by the high variation in immunity within the population to SARS-CoV-2 infection and COVID-19 severity.

As a consequence of population heterogeneity, it is no-longer possible or reasonable to utilise a simply defined effectiveness that relates the protection offered by a COVID-19 vaccine to the risks experienced by unvaccinated individuals [7]. The analysis of the potential impact and cost-effectiveness of future COVID-19 vaccination programmes therefore requires a pragmatic quantitative approach that accounts for the extreme heterogeneity in population immunity. To assess the impact and cost-effectiveness of prospective routine COVID-19 vaccination programmes for England, in this study we take the simplifying approach of contrasting individuals who have recently been vaccinated (in the last six months) with those that have not.

We use COVID-19 epidemiological data from England for March 2023 to March 2024, that also partitions the population by age and by three risk groups (not at risk, at risk but not immunosuppressed, and immunosuppressed). We present a collection of methodological approaches (each with differing assumptions) to calculate the risk of severe outcomes (hospital admission, ICU admission or death) and the protection offered by COVID-19 vaccination. We combine these estimates with bespoke health economic assessments that capture age and risk dependent heterogeneity in hospital cost and QALY (quality adjusted life years) losses following admission or death. This analysis allows us to calculate the willingness to pay threshold, which determines the price at which costs and benefits are equal.

## 2. Methods

### Study population

Throughout our analysis, we stratified the adult population into 16 different five-year age cohorts (*a*) from 15-19 to 90+, and three different risk groups (*r*): not at risk; at-risk (but not immunosuppressed); and immunosuppressed. The immunosuppressed group accounts for only 1.7% of the English population (peaking at 4.9% of those 80-84 years old), and therefore any results are subject to considerable uncertainty; we hence restrict our attention to the other two risk stratifications but return to the immunosuppressed group in the discussion.

For each of the sub-groups we had data for the population size, the number of hospital admissions, severe hospital admissions (for brevity we refer to as ICU admissions) and deaths each week partitioned between those vaccinated in the last six months and those unvaccinated in that same time period (Fig. 1). This data allowed us to calculate the rates (confidence intervals and full distributions) of different severe health outcomes for each of the sub-groups. If the uptake of vaccine is assumed to be random within a sub-group, then the ratios between the rates associated with recently-vaccinated and not recently-vaccinated groups defines the realised vaccine effectiveness. This realised vaccine effectiveness should account for the heterogeneous levels of pre-existing partial immunity in the population, including the impacts of past infection and past vaccination. However, it is important to consider some caveats with these data, chiefly that: the severe outcomes data does not separate infection episodes where COVID-19 was the primary or secondary cause; and that we assume outcomes are random within an age- and risk-group (i.e. there is no correlation between the proportion that get vaccinated, take additional precautions or are more severely affected). In the Discussion we elaborate on the potential implications of these data caveats on our findings.

**Figure 1.**
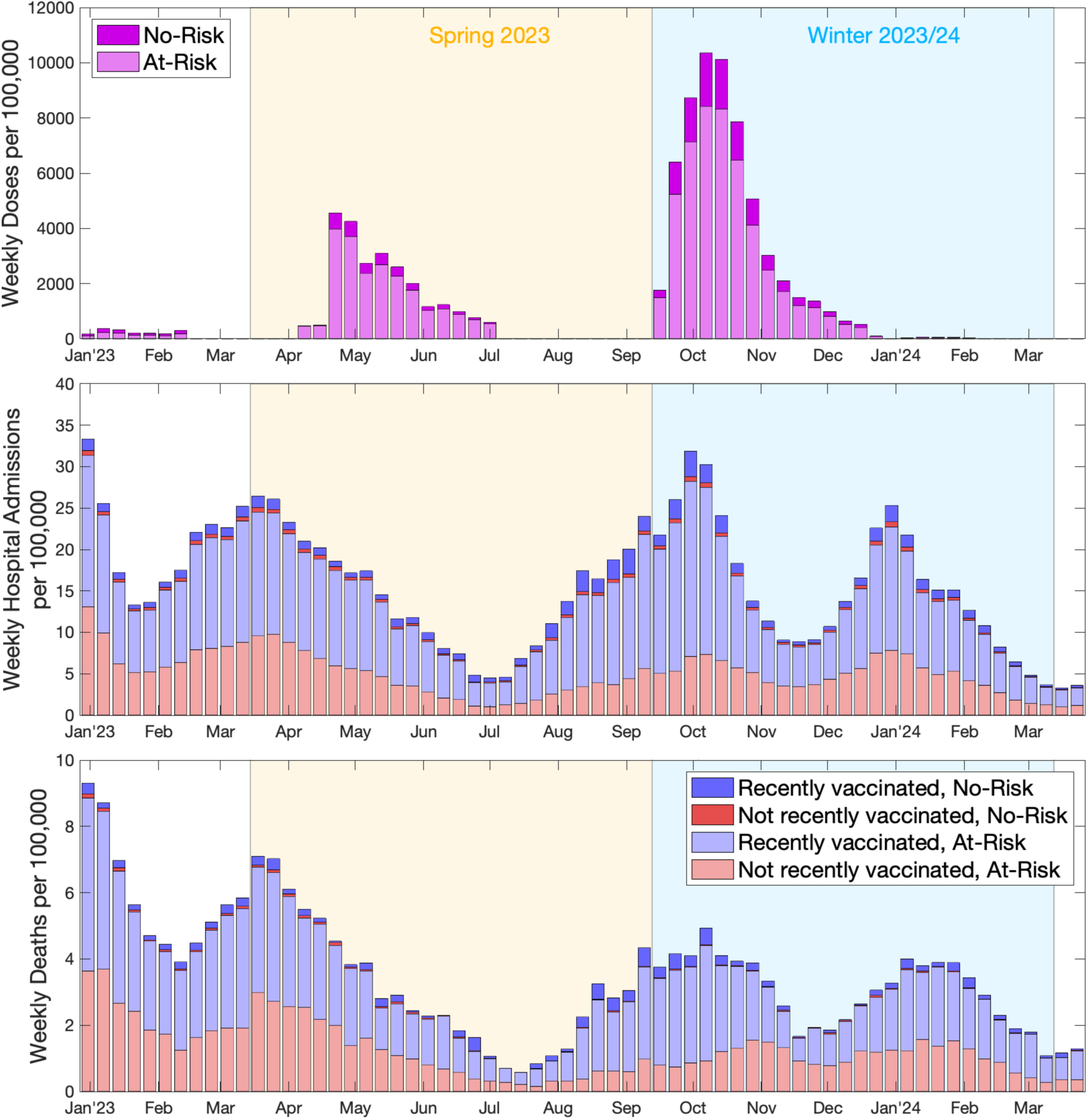
COVID-19 vaccination and severe outcomes data from 2023 to 2024 in England. We display per week and per 100,000 individuals: vaccinations administered (top); number of hospital admissions (middle); number of deaths (bottom). The data is partitioned by risk group (no-risk in darker shades and at-risk in lighter shades) and vaccination status (recently vaccinated in blue and not recently vaccinated in red). The time period is divided into two 26-week seasons (Spring 2023 in orange and Winter 2023/24 in blue), which capture the spring 2023 or autumn 2023 booster campaigns and the major epidemic waves. The data come from the Secondary Uses Service [8], the ONS death records [9] and UKHSA’s Immunisation Information System [10].

### Study time periods

For the purpose of this study, we partitioned the data into two equal time intervals of 26 weeks associated with protection from the spring and autumn 2023 booster campaigns: spring/summer 2023, from week 11 to week 36 of 2023 (roughly mid-March to mid-September); and winter 2023/24, from week 37 of 2023 to week 10 of 2024 (roughly mid-September to mid-March the subsequent year).

We review the temporal pattern of vaccination and severe outcomes (hospital admissions and deaths), summing across all ages, but differentiating by risk-group and vaccination status (Fig. 1). The number of vaccine doses administered (Fig. 1, top panel) has two clear periods of activity (Spring and Autumn); this contrasts with the weekly hospital admissions (1, middle panel) and weekly deaths (Fig. 1, bottom panel), which do not show such clear seasonal patterns.

Given the lack of seasonal structure in the epidemiological data we acknowledge the 26-week time partitions are somewhat arbitrary. Nonetheless, the results are generally insensitive to the precise temporal divisions.

### Vaccine effectiveness methodologies

We now describe the statistical analysis of these data using the three methodologies (providing five distinct effectiveness estimates). The first method we call the ‘aggregate approach’, the second method we call the ‘time since vaccination approach’ and the third method we call the ‘literature-based vaccination effectiveness’.

For all three methodologies we computed joint probability distributions for the rate of severe disease of recently vaccinated and not recently vaccinated individuals (alternatively, this is equivalent to the joint distribution on the rate in not recently vaccinated and the effectiveness of vaccination). We independently calculated joint distributions for the three measures of severe disease (hospital admission, ICU admission/severe hospitalised cases, and deaths), for the different risk groups, the two seasons (associated with the spring and autumn booster) and, applicable only to our aggregate approach (Method 1), for each age group. We summarise below each methodology, which are described mathematically in the Supplementary Methods.

#### Aggregate approach, Method 1

The simplest approach used aggregate data from each season, risk-group and age-group to inform the risks of severe disease in the recently vaccinated and not recently vaccinated populations. As such, we averaged all population sizes and rates over the 26-week seasons. We could carry out this calculation for all age and risk groups where vaccination was generally available; for the non-risk group this was those aged 75 years and above in Spring 2023 and those aged 65 years and above in Autumn 2023.

#### Time since vaccination approach, Method 2

This more sophisticated methodology includes three important factors: the weekly pattern of severe disease and vaccination; the waning of protection over time; and extending the results to the entire population. We implemented this extension to the entire population by calculating a single effectiveness probability distribution for all ages (although note we calculated different distributions for each season and risk-group); this was based on the ages where vaccination was generally available and then extended to younger age-groups. We modelled the waning of vaccination projection in two ways. In Method 2a, we generated the average population level of protection (for those vaccinated in the last six-months) each week, using an assumed level of waning that matched observations from the Delta and Omicron (BA.1) waves. In Method 2b, we estimated the decline in protection to maximise the likelihood. Throughout this calculation the likelihood was computed using the weekly data.

#### Literature-based vaccine effectiveness, Method 3

Finally, effectiveness values and levels of protection against Omicron have been calculated by multiple other groups [11,12], with estimates from the UK providing high-quality and early estimates [13]. Here we use two recent estimates (at the time of writing) from the UK: (i) estimates produced by Public Health Scotland measured between 04 September 2023 and 31 March 2024 [14]; (ii) estimate produced by the UK Health Security Agency measured between 04 September 2023 and 21 January 2024 [15]. These estimates gave a distribution of effectiveness values (and their decline) that we incorporated into the joint distribution estimate of rates of severe disease. As with Method 2, we used the same effectiveness values for all age-groups.

### Willingness to pay thresholds

By combining our estimates for the rates of severe outcomes and the level of vaccine protection with the health economic estimates from a bespoke study, we arrive at a willingness to pay threshold for the total cost of the vaccine and administration. We followed the guidelines established by the Joint Committee on Vaccination and Immunisation (JCVI - an expert scientific advisory committee which advises the UK government on vaccination and immunisation matters) and calculated two quantities: a central estimate of the willingness to pay threshold (using the maximum likelihood estimates and valuing one Quality Adjusted Life Year or QALY at £20,000) and the threshold for which 90% of scenarios are cost-effective (using £30,000 per QALY and fully accounting for uncertainty). The final willingness to pay threshold was the minimum of these two values, which for the vaccination programmes considered here was generally the central estimate [16].

## 3. Results

### Aggregate approach (Method 1)

The simplest method (Method 1) used the aggregate data from one of the 26-week long time periods of interest (the Supplementary Methods section provides a detailed mathematical description). In principle, this approach can provide estimates of vaccine effectiveness for each age and risk group. However, the results are not necessarily reliable when the uptake is biased. For example, during the Autumn 2023 booster programme everyone 65 years of age and above was eligible; for those under 65 but not in a risk group about 1.5 million doses were administered, often due to them being carers or working in frontline health and social care - such individuals may have greater exposure to infection than others of the same age, which would lead to biases in vaccine effectiveness estimates. For this reason, we only applied this first method to age-groups where there was a universal offer of vaccination (75 years and above for Spring 2023; 65 years and above for Autumn 2023 booster). As such, Method 1 generates an effectiveness that depends on age, season, and risk-group; although the protection does not decline with time-since vaccination. As an exemplar of the calculations conducted for this method, we consider those not at risk and aged 80-84 during the winter 2023/24 season, and calculate the central estimate of the willingness to pay threshold.

There were 412,436 individuals in this age and risk group, of whom an average of 335,928 had been vaccinated in the previous six months – leaving 76,508 not recently vaccinated. In the recently vaccinated group there were 186 hospital admissions (a rate of 55.4 per 100,000 over the six-month period), while in the not recently vaccinated group there were 128 hospital admissions (a rate of 167.3 per 100,000). Combining these numbers returned a vaccine effectiveness of 66.9% against hospital admission.

In addition, those that have been recently vaccinated had a shorter average length of stay in hospital: 12.8 days compared to 15.0 days for those not recently vaccinated. The average rates and lengths of stay mean that for every thousand doses of vaccine administered to this age and risk group, 1.12 hospital admissions and 18.0 days of hospital stay are prevented. (the 18.0 days saving are comprised of both reduced admissions and shorter stays.) These savings can be combined with the health economic estimates that every admission for this age and risk group is associated with an average 0.299 QALY loss, and that each day in hospital costs an average of £1182, to produce a willingness to pay threshold of £27.98 (= [1.12 × 0.299 × £20,000 + 18.0 × £1182]/1000) per dose for its protection against hospital admission alone.

### Time since vaccination approach (Method 2)

Our second approach (Estimate 2a) used an assumed decline in protection over time (dropping by 74% over six months), generating a population-level average degree of protection (relative to a recently vaccinated individual) for each week. This again allowed us to estimate the basic rates of severe outcomes and the associated vaccine effectiveness, and hence to determine the total amount of protection offered by a vaccine over the six-month period. To overcome the issues of biased vaccination in those under 65 year of age in the no-risk group (as mentioned above), we extrapolate the vaccine effectiveness estimates for all no-risk individuals over 65 to the entire no-risk population. We perform a similar extrapolation for the at-risk population, using the effectiveness estimated from at-risk individuals over 65 years of age. As such, Method 2 generates an effectiveness that depends on season and risk-group (but not age), where the protection declines with time-since vaccination.

Considering hospital admissions, our central estimate of the vaccinate effectiveness (in the no-risk group) was 70.0% immediately after vaccination, with an average vaccine effectiveness of 47.0% across a six-month period. For not-at-risk 80-84 year olds, this adjusts the previous estimates such that for every thousand doses of vaccine, 0.606 hospital admissions and 10.7 days of hospital stay are prevented. Combining these numbers with the health economic costs generated a willingness to pay threshold of £16.27 per dose for its protection against hospital admission.

As an extension to this approach, Estimate 2b used a maximum likelihood approach to determine the appropriate decline in protection since vaccination for hospitalisation and death outcomes separately, based on the data for those over 65 years of age. These estimates suggest a slower decline in protection than assumed for Estimate 2a, and are comparable with recently estimates from Scottish data [14]. For hospital admissions in the no-risk group, our revised central estimate of vaccine effectiveness was 62.8% immediately after vaccination, with an average vaccine effectiveness of 57.7% across a six month period. These revised protection values, change our central estimates for no-risk 80-84 year olds to preventing 0.639 hospital admissions and 11.4 days of hospital stay per thousand doses of vaccine; leading to a willingness to pay threshold of £17.24 per dose for protection against hospital admission.

### Literature-based vaccine effectiveness (Method 3)

Our third and final method used vaccine effectiveness estimates derived from alternative data sets. The approach could be used for any set of vaccine effectiveness estimates [17]; here we make use of two UK-based estimates: Estimate 3a uses vaccine effectiveness and waning of protection from Public Health Scotland (PHS) [14]; Estimate 3b uses effectiveness estimates from UKHSA [15]. The PHS estimates come from individuals aged over 65 that were part of the Evaluation and Enhanced Surveillance of COVID-19 (EAVE-II) cohort, and included a surveillance period of September 2023 to July 2024. The UKHSA estimates used a test-negative case-control study design, where positive PCR tests from hospitalised individuals are cases and comparable negative PCR tests are controls; this approach has been used to produce official vaccine estimates for the UK [18, 19]. Our approach utilises a probability distribution for the effectiveness which is then scaled in a deterministic manner over time. We use this formulation to capture the uncertainty in the peak level of protection, and the pattern of change over time follows the mean of the PHS or UKHSA estimates. As such, Method 3 generates an effectiveness that is the same for all ages, seasons and risk groups although the protection does decline with time-since vaccination.

For hospital admissions, the PHS estimates generated a mean vaccine protection across a six-month period of 65.3% (Method 3a), while the UKHSA estimates led to 27.1% protection (Method 3b). For not-at-risk 80-84 year olds, these vaccine effectiveness estimates correspond to willingness to pay thresholds of £18.74 and £8.86 per dose respectively for protection against hospital admission.

### 3.1 Willingness to pay thresholds

Similar calculations can be performed for ICU admissions and deaths, for different age and risk groups and for Spring 2023 and Autumn 2023 boosters; the full calculation also computes the probability distribution associated with the rates and therefore captures the uncertainty in the vaccine protection (see Supplementary Methods). These statistical results can then be combined with the health economic costs and benefits, which also capture uncertainties to generate a total willingness to pay threshold accounting for all the outcomes and following the JCVI guidelines on dealing with uncertainty.

For the spring 2023 and autumn 2023 boosters, and the six-month period of COVID-19 infection, Fig. 3 shows the willingness to pay threshold for different ages, two different risk groups and the five different estimates for the rate and effectiveness distributions. We did not apply Method 1 (aggregated calculation method) to groups with insufficient data or where we believed there was a notable bias between those receiving a booster dose and those that did not (under 65 for the autumn 2023 booster and under 75 for the spring 2023 booster). For each Method, we independently calculated vaccine effectiveness values for each booster season (spring 2023, autumn 2023) and for protection against the three forms of severe disease outcome (hospital admission, ICU admission and death).

All estimates, risk-groups and seasons show the same basic trend: willingness to pay increases dramatically with age, capturing the well-recognised observation that older individuals are more at risk of severe outcomes following COVID-19 infection [20, 21, 6]. The results also highlight the generally higher willingness to pay (across all age-groups and estimates) for individuals classified as at-risk compared to no-risk, due to the higher chance of more severe outcomes. Finally, we observe higher willingness to pay for the autumn 2023 booster compared to the spring 2023 booster, due to the higher number of infections in the associated winter period. We note that the JCVI require both the central estimate (at £20,000 per QALY) and 90% of uncertainty scenarios (at £30,000 per QALY) to be cost effective - this translates to the final willingness to pay threshold being the minimum of the most likely value (dot in Fig. 3) and 10^th^ percentile of the estimated willingness to pay threshold distribution (lower bounds of the narrower boxes in Fig. 3).

We consider the implications for the willingness to pay threshold for each age-group (Table 1) in terms of who should be offered the vaccine. Given the uncertainty in the seasonality of COVID-19 cases (we have yet to experience a single distinct winter wave or even a consistent seasonal pattern) we amalgamate results from the spring 2023 and autumn 2023 boosters, and report the average willingness to pay threshold across both boosters. In addition, it is thought that there is substantially better vaccine uptake from a universal programme rather than one that targets at-risk groups. We therefore also report the weighted average, which gives the willingness to pay threshold for a universal programme. We again see the general trends that older age-groups have a higher willingness to pay threshold, such that vaccination is more likely to be cost-effective.

**Table 1:** Willingness to pay thresholds for the combined Spring/Autumn 2023 boosters. . Thresholds are given for all ages (although ages 15-39 have been amalgamated for simplicity), for no-risk, at-risk and universal vaccine deployment and for the five effectiveness estimates. Values are rounded to the nearest pound, and values less than £1 are indicated. Ages, risk-groups and methods where the willingness to pay threshold is determined by the JCVI uncertainty condition (that there is a 90% chance of cost-effectiveness at £30,000 per QALY) are marked with an asterisk.

|  | Universal |  |  |  |  | No Risk |  |  |  |  | At Risk |  |  |  |  |
| --- | --- | --- | --- | --- | --- | --- | --- | --- | --- | --- | --- | --- | --- | --- | --- |
| Age | E1 | E2a | E2b | E3a | E3b | E1 | E2a | E2b | E3a | E3b | E1 | E2a | E2b | E3a | E3b |
| 15-39 | - | < £1 | < £1 | < £1 | < £1 | - | < £1 | < £1 | < £1 | < £1 | - | ≤ £2 * | ≤ £2 * | ≤ £3 | ≤ £2 * |
| 40-44 | - | £1 | £1 | £1 | < £1 | - | < £1 | < £1 * | < £1 | < £1 | - | £3 | £2 | £5 | £2 |
| 45-49 | - | £1 | £1 | £2 | £1 | - | £1 * | < £1 | £1 | < £1 | - | £3 | £3 | £6 | £3 |
| 50-54 | - | £2 | £2 | £3 | £2 | - | £1 | £1 | £1 | £1 | < £1 | £6 | £5 | £10 | £5 |
| 55-59 | - | £3 | £3 | £5 | £2 | - | £1 | £1 | £1 | £1 | < £1 | £8 | £7 | £13 | £6 |
| 60-64 | - | £5 | £4 | £7 | £4 | - | £2 | £2 | £2 | £1 | £2 * | £10 | £8 | £16 | £8 |
| 65-69 | £8 * | £11 | £9 | £16 | £8 | £4 * | £4 | £3 | £4 | £2 | £14 * | £19 | £16 | £30 | £15 |
| 70-74 | £17 | £16 | £13 | £25 | £12 | £8 * | £7 | £6 | £7 | £4 | £25 | £24 | £20 | £41 | £19 |
| 75-79 | £51 | £33 | £30 | £57 | £24 | £31 | £17 | £17 | £19 | £8 | £63 | £44 | £38 | £81 | £33 |
| 80-84 | £59 | £46 | £41 | £89 | £35 | £37 | £27 | £27 | £31 | £13 | £67 | £53 | £46 | £112 | £44 |
| 85-89 | £103 | £86 | £75 | £160 | £64 | £72 | £58 | £60 | £71 | £30 | £111 | £93 | £80 | £183 | £72 |
| 90+ | £85 | £89 | £77 | £188 | £72 | <<br>£1 | £64 | £63 | £77 | £36 | £103 | £94 | £80 | £210 | £79 |

### Pre-purchased vaccine

The COVID-19 booster vaccines used during spring and autumn 2023 were pre-purchased during the pandemic; they can therefore be considered to have zero price. The only cost of vaccination is therefore the administration costs, which have generally been between £7.50 and £10.04 (although higher values may apply for administration to house-bound individuals). Using this range as our cost, we observe that vaccination of no-risk individuals aged 75 and older is likely to be cost-effective for all methods of determining effectiveness, although for Estimate 3b (UKHSA vaccine effectiveness [15]) this will be sensitive to the precise costs of administration. We note that even a vaccine with 100% effectiveness (not shown) is not cost effective for no-risk individuals under age 65 as the administration costs are greater than the benefits. For at-risk individuals, the age at which vaccination is cost effective reduces to either 60 or 65 years of age depending on the method used, while for a universal vaccination programme the age threshold is either 65 or 70 years old.

Taking Method 2 (Estimates 2a and 2b) as our benchmark, we would therefore suggest that the ideal strategy for a pre-purchased vaccine would be to offer a universal vaccination to all individuals over 70, and also to offer vaccination to at-risk individuals over 65. While vaccination of those aged 70-74 in the no-risk group is not predicted to be cost-effective (willingness to pay is £6-7), the advantages in terms of higher uptake in the at-risk group of a universal programme is likely to off-set any costs in the no-risk group.

If we consider the spring 2023 and autumn 2023 boosters separately then we find the age thresholds for universal vaccination are 75 and 65 respectively (Tables S1 and S2 in the Supplementary Material). These estimates come with the benefit of hindsight, such that the scale of the outbreak is known, as is the vaccine effectiveness; therefore it is remarkable that these are in agreement with the JCVI advice for individuals not at risk for these booster programmes [22, 23].

### Purchased vaccine

In the immediate future, it will become necessary for the UK to purchase vaccines and place vaccination against COVID-19 on a equal cost-benefit assessment as all other vaccines. Which age-groups it is cost-effective to vaccinate will be highly dependent on the price paid for the vaccine, which is currently unknown. To illustrate the patterns, here we consider £25, £50 and £75 as combined costs for the vaccine and administration.

At £25 per dose delivered, all Methods broadly agree that universal vaccination of those aged over 75 is likely to be cost effective, assuming that losses from vaccinating no-risk individuals aged 75-79 are compensated for by an increased uptake in at-risk individuals in the same age-group. It is uncertain whether vaccination of 70-74 year olds in the at-risk group will be cost-effective with different effectiveness estimates producing conflicting results, potentially suggesting a need to refine the co-morbidities that define at-risk.

At £50 per dose delivered, the age threshold for universal vaccination increases to 80 (Estimates 1 and 3a) or 85 (Estimates 2a, 2b and 3b). Only estimates 1 and 3a suggest that vaccinating younger individuals (those aged 75-79) in at-risk groups could be cost-effective. Finally, at £75 per administered dose, it is likely that universal vaccination is only cost-effective for those over 85; although Method 3a suggests universal vaccination is not cost-effective at any age, and only vaccination of at-risk individuals over 90 is cost-effective.

### 3.2 Sensitivity

Some of the uncertainties in our epidemiological and health-economic model can be quantified. For example, based on the available data, inferring the rate of hospital admissions produces a detailed distribution of parameter uncertainty (Fig. 2). However, for other elements - such as those associated with the precise scale and timing of future waves, or the degree of protection against future variants - the uncertainty cannot be quantified; for example, we cannot determine the probability that the 2024/25 winter wave will be bigger (or smaller) than the 2023/24 winter wave. For these uncertainties we present an uncertainty analysis, showing the willingness to pay threshold as a function of the unknown quantity.

**Figure 2:**
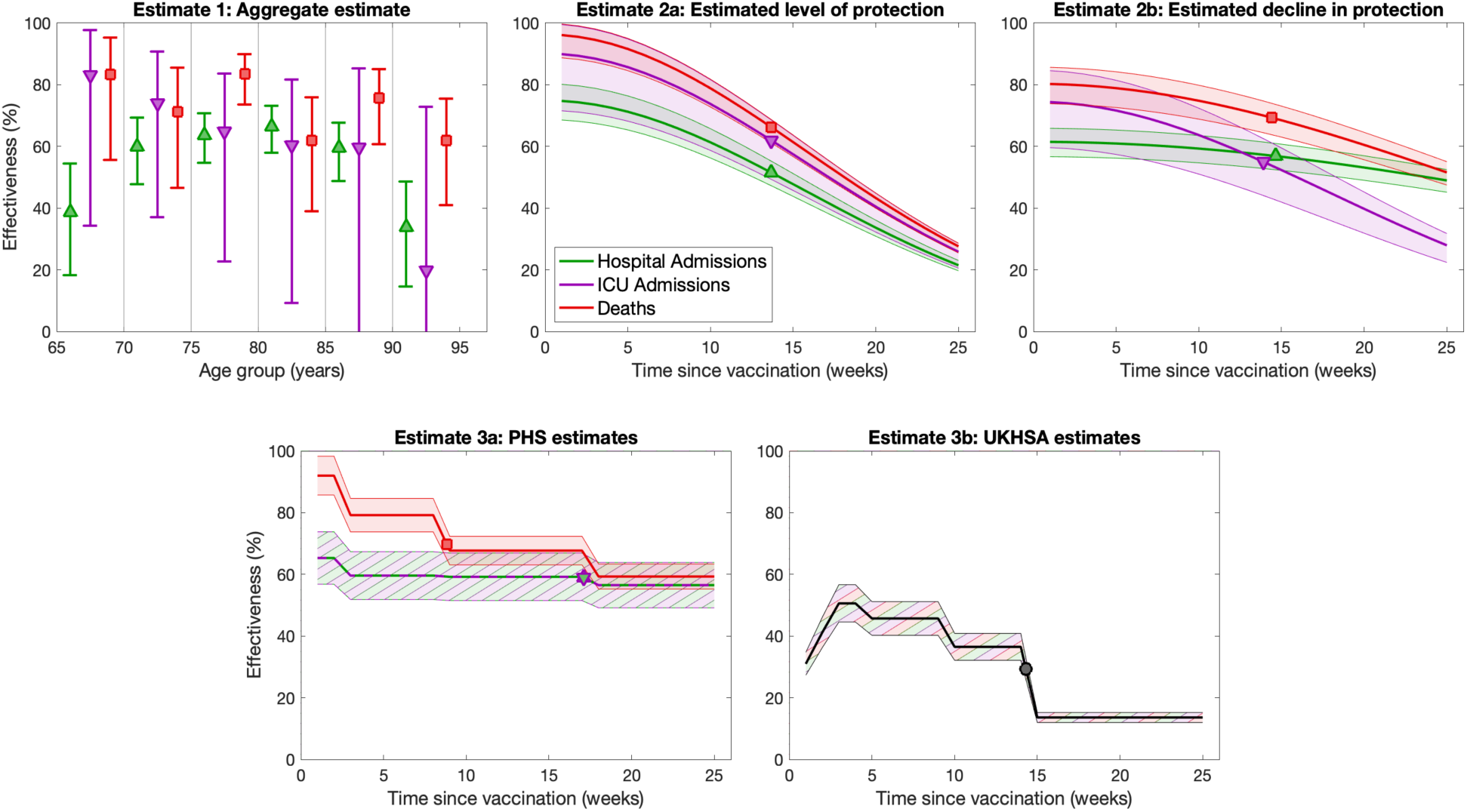
Effectiveness estimates from the five approaches, for no-risk individuals following the autumn 2023 booster. Throughout we present effectiveness estimates for protection against hospital admissions in green (triangle symbol), protection against ICU admissions in purple (inverted triangle) and protection against mortality in red (square). For the aggregate estimate (Estimate 1), where we assumed vaccine effectiveness was fixed across a six-month interval, we estimated the effectiveness for each 5-year age group that was universally offered the vaccine. We display the 95% prediction intervals (spanning 2.5th and 97.5th percentile), with symbols denoting the mean values. Estimates 2a and 2b show the inferred effectiveness distributions with either an assumed or fitted decline. Estimates 3a and 3b are based on values from PHS [14] and UKHSA [15]; the PHS estimates assumed the same effectiveness for hospital and ICU admission, while the UKHSA estimates assumed the same effectiveness for all severe outcomes. Throughout we show 95th percentiles of the prediction interval (shaded ribbons) as well as the mean value (solid lines); symbols show the mean level of effectiveness over a six-month period.

**Figure 3:**
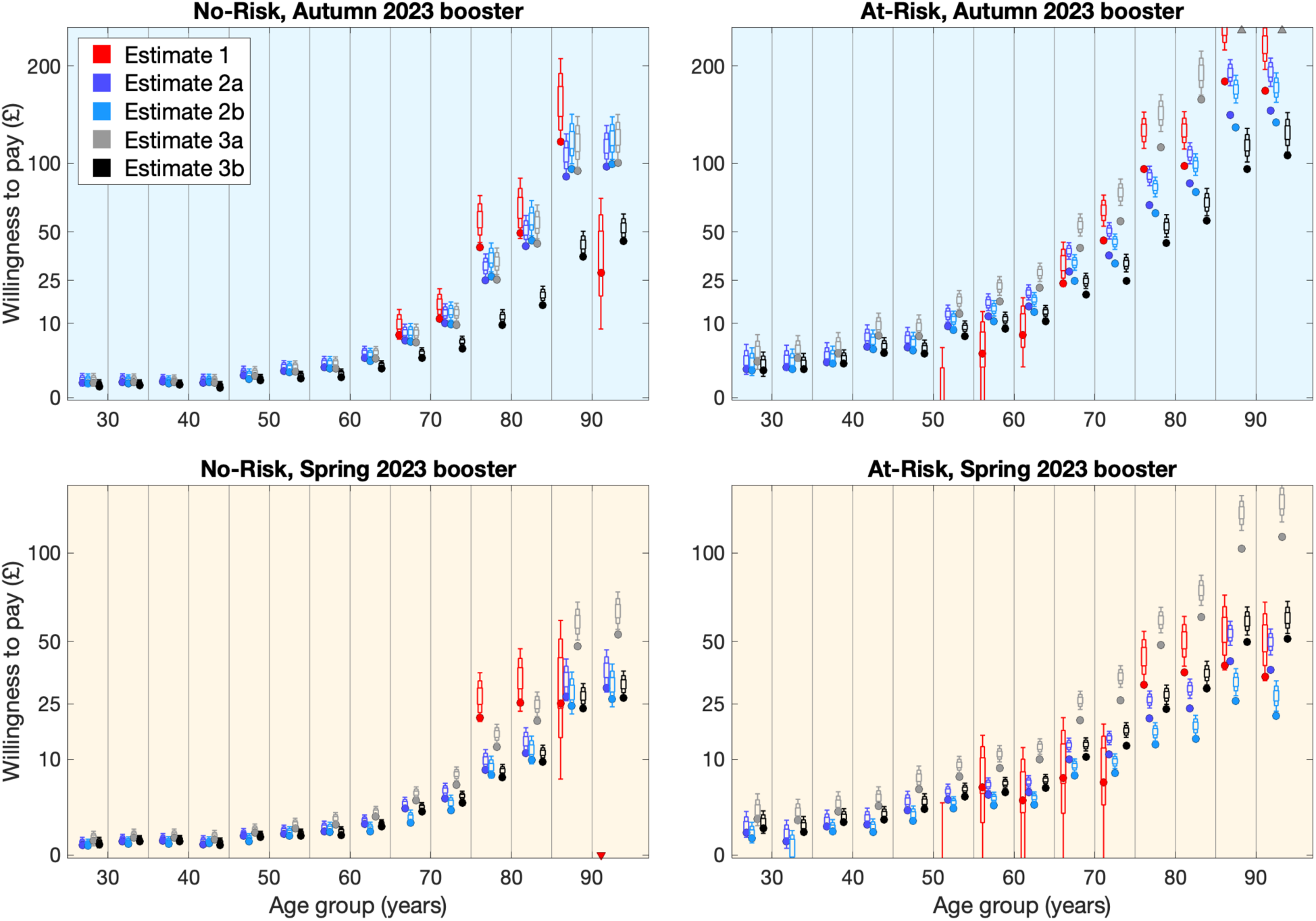
Using data from the spring and autumn 2023 boosters and the 2023/24 winter wave of COVID-19, vaccine threshold prices stratified by age-group, risk-group and modelling approach. We calculate the vaccine threshold price for the two time periods (top row: winter 2023/24; bottom row: spring 2023), 16 age groups (y-axis), three risk groups (panels) and five methodologies (red, dark-blue, light-blue, grey and black). Dots show the most likely value assuming £20,000 per QALY; when assuming £30,000 per QALY the extended bar-and-whisker plots show the 95%, 80% and 50% credible intervals combining all sources of uncertainty. Points that are above or below the axis are shown as triangles. Note that the y-scale is non-linear to better display lower willingness to pay thresholds.

Retaining our focus on universal vaccination, and using Method 2 (Estimates 2a and 2b) throughout, we consider three sensitivities: a shift to the timing of the wave(s); a rescaling of the size of the wave(s); and different levels of vaccine protection (Fig. 4). We also use a combined cost of £25 per administered dose to illustrate the impact of these uncertainties, but the implication of other costs can be assessed from Figure 4.

**Figure 4:**
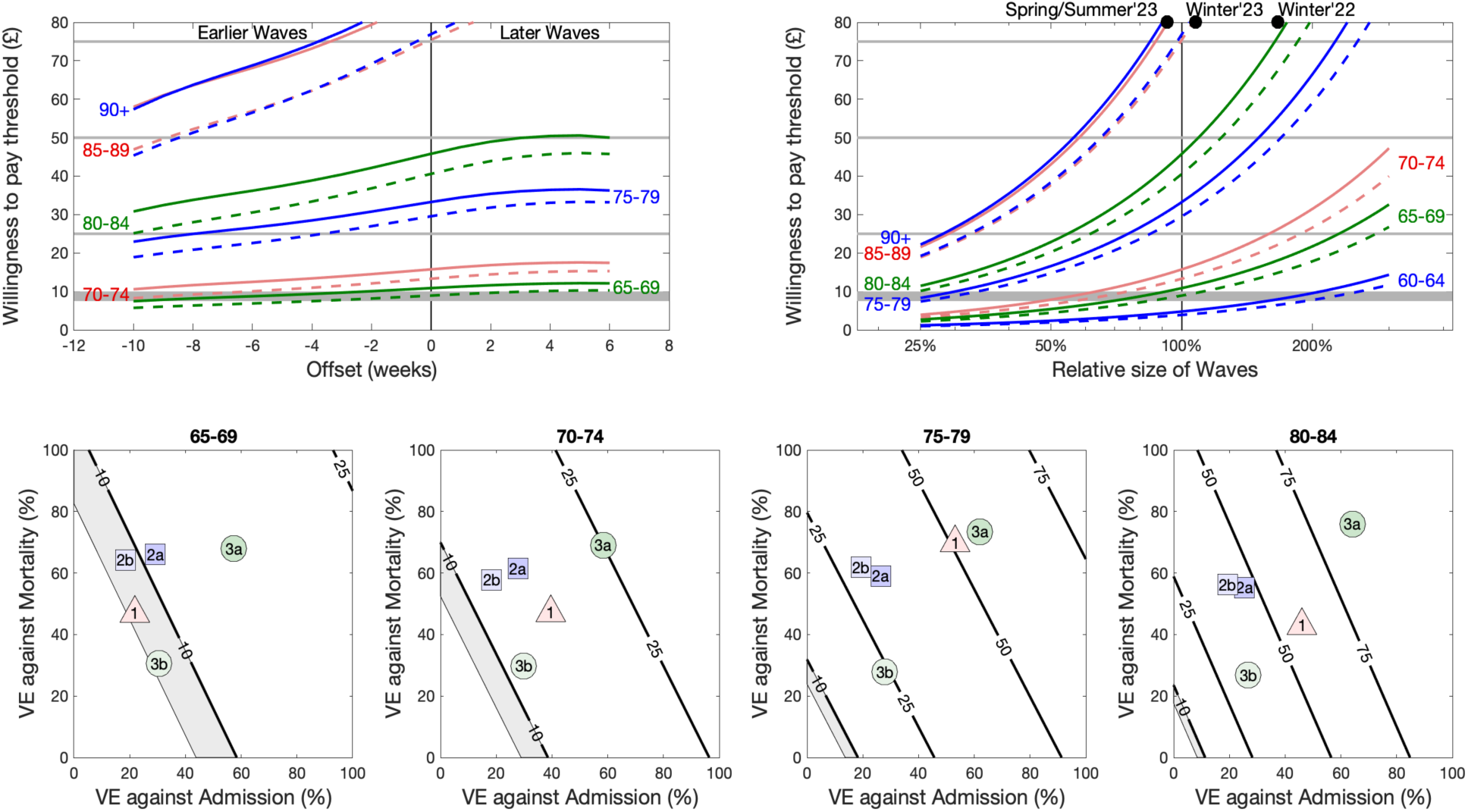
Sensitivity of the willingness to pay threshold for universal vaccination to the timing of the epidemic wave, the magnitude of the epidemic waves and the effectiveness of the vaccine. For a shift in the timing of waves or waves of different sizes (top row), we consider how the threshold calculated by Estimates 2a (solid line) and 2b (dashed line) for older age-groups vary. When considering the size of the waves, the Spring 2023, Winter 2023-24 and Winter 2022-23 are shown for comparison. For vaccine effectiveness (VE), we extrapolate the willingness to pay thresholds from Estimate 2a and consider protection against hospital admissions and mortality separately (bottom row). We consider four age-groups (65-69 to 80-84) and show the contours where the willingness to pay threshold is equal to £7.50-£10.00 (grey shaded region), £25, £50 and £75; we also show the VE estimates for the five approaches, weighted so that they apply to universal vaccination over combined spring 2023 and autumn 2023 boosters.

Changes to the timing of the waves has the smallest impact on the willingness to pay thresholds (Fig. 4, top-left). Earlier waves consistently reduce the threshold, such that if the waves were 4-7 weeks earlier (depending on the Method) it may no-longer be cost effective to offer universal vaccination to those aged 75-79. There is some evidence that earlier vaccination by around four weeks could increase the benefit of vaccination (assuming the timing of the waves remained the same).

The size of the waves of hospital admissions and deaths has a linear impact on the willingness to pay (Fig. 4 top-right). At the £25 cost, very large waves could lead to vaccination of 65-69 year olds being cost effective, although this would require outbreaks more severe than those of Winter 2022-23. Similarly, waves that are consistently 15-25% smaller lead to the vaccination of 75-79 year olds no-longer being cost-effective.

We consider uncertainty in vaccine effectiveness to capture either improvements in vaccine targeting or the risk of evolution of vaccine escape. For the 65-69 age group, it would require a near perfect effectiveness for the vaccination of this age-group (at a cost of £25) to be cost effective. Similarly, a large improvement in effectiveness is required for universal vaccination of those aged 70-74 to be cost effective at £25 per administered dose. For older age-groups (for example 80-84 year olds), the effectiveness would need to be extremely low to change the cost effectiveness.

## Discussion

Here we have quantified the protection offered by recent COVID-19 booster vaccination by using national data sets on COVID-19 associated hospital admissions, ICU admissions and deaths in England linked to individual vaccination records. We used three different approaches leading to five methods to infer the underlying risk of severe outcomes in different age and risk groups (no-risk, at-risk or immunosuppressed, as defined in the Green book chapter 14a [24]) dependent on vaccination status. We combined these inferred risks with bespoke health economic parameters for COVID-19 on hospital costs and QALY losses to define willingness to pay thresholds according to JCVI guidelines. As such, the willingness to pay threshold defines the maximal combined cost of a vaccine dose plus the administration, such that vaccination would be cost effective; the JCVI guidelines ensure that both the most likely scenario and uncertainty are taken into account in this evaluation.

The willingness to pay threshold we calculate is used to translate the total vaccination cost (vaccine plus administrative costs) into a set of age and risk groups who it should be cost effective to vaccinate. For COVID-19, given the increasing vulnerability with age, this equates to a lower age threshold in each risk group, with everyone older being eligible for vaccination. Table 2 shows this lower age threshold for a range of costs and for universal vaccination and at-risk groups; based on the assumption the universal vaccination has the benefit of increased uptake in the at-risk population). This analysis assumes that the same threshold is applied to both spring and autumn boosters, given the absence of a clear seasonal pattern in COVID-19 cases and outcomes to date. For each cost we show the lowest and highest estimated age threshold together with the associated models. Despite the very different assumptions underpinning the five estimates, there is a remarkable level of agreement, in part due to the extreme increase in risk with increasing age. We find that Estimate 3a (PHS estimates) is always associated with the lowest age estimates, while Estimate 3b (UKHSA effectiveness estimates) is always associated with the highest estimates - providing bounds on who should be considered for cost-effective vaccination.

**Table 2:**
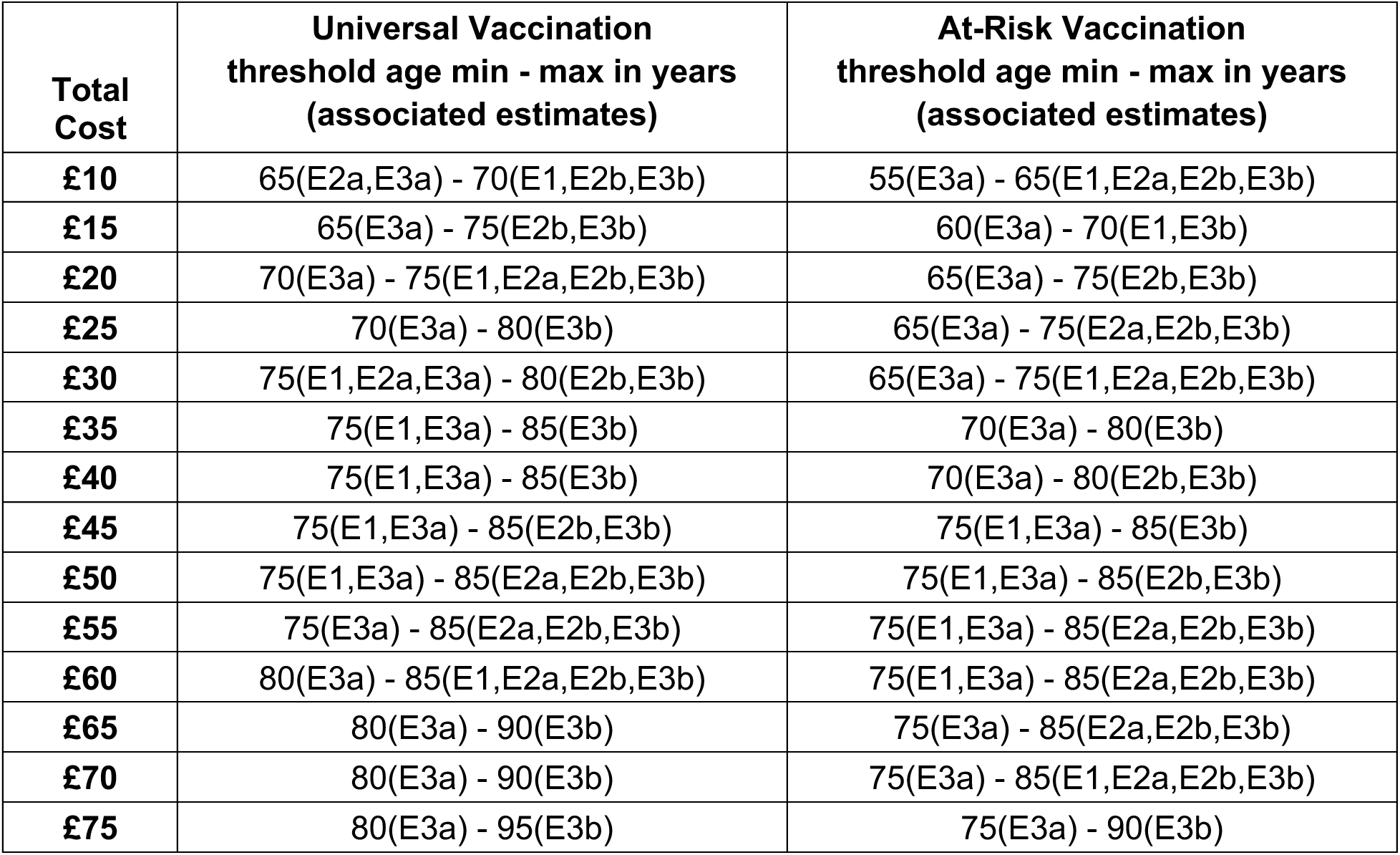
The lower age threshold for cost-effective vaccination using the imputed willingness to pay values. For each combined cost of vaccine and administration, we give the minimum and maximum calculated age thresholds above which vaccination is cost effective, together with the estimates that generate these thresholds.

| **Total Cost** | **Universal Vaccination**<br>**threshold age min - max in years**<br>**(associated estimates)** | **At-Risk Vaccination**<br>**threshold age min - max in years**<br>**(associated estimates)** |
| --- | --- | --- |
| **£10** | 65(E2a,E3a) - 70(E1,E2b,E3b) | 55(E3a) - 65(E1,E2a,E2b,E3b) |
| **£15** | 65(E3a) - 75(E2b,E3b) | 60(E3a) - 70(E1,E3b) |
| **£20** | 70(E3a) - 75(E1,E2a,E2b,E3b) | 65(E3a) - 75(E2b,E3b) |
| **£25** | 70(E3a) - 80(E3b) | 65(E3a) - 75(E2a,E2b,E3b) |
| **£30** | 75(E1,E2a,E3a) - 80(E2b,E3b) | 65(E3a) - 75(E1,E2a,E2b,E3b) |
| **£35** | 75(E1,E3a) - 85(E3b) | 70(E3a) - 80(E3b) |
| **£40** | 75(E1,E3a) - 85(E3b) | 70(E3a) - 80(E2b,E3b) |
| **£45** | 75(E1,E3a) - 85(E2b,E3b) | 75(E1,E3a) - 85(E3b) |
| **£50** | 75(E1,E3a) - 85(E2a,E2b,E3b) | 75(E1,E3a) - 85(E2b,E3b) |
| **£55** | 75(E3a) - 85(E2a,E2b,E3b) | 75(E1,E3a) - 85(E2a,E2b,E3b) |
| **£60** | 80(E3a) - 85(E1,E2a,E2b,E3b) | 75(E1,E3a) - 85(E2a,E2b,E3b) |
| **£65** | 80(E3a) - 90(E3b) | 75(E3a) - 85(E2a,E2b,E3b) |
| **£70** | 80(E3a) - 90(E3b) | 75(E3a) - 85(E1,E2a,E2b,E3b) |
| **£75** | 80(E3a) - 95(E3b) | 75(E3a) - 90(E3b) |

For low total costs (generally associated with the assumption that the vaccine is pre-purchased and therefore zero cost), we find that universal vaccination is cost-effective for those aged over either 65 or 70 depending on the methodology employed; however, the majority of the models agree that for those in the at-risk group, vaccination should be offered to those 65 and over. For slightly higher combined costs, for example at £25, most methods (Estimates 1, 2a and 2b) agree that the universal threshold should be those over age 75, and this is also the most preferred age threshold for the at-risk group. For much higher costs, for example at £50-60, the majority of the models suggest that universal vaccination should only be offered to those over the age of 85, with weak or no support for vaccination of younger at-risk individuals.

Those who are immunosuppressed form a small but highly vulnerable part of the population. Due to the small size of this risk group, and the heterogeneity within it, results from all estimations of effectiveness are extremely noisy. However, as a general rule, vaccination of all immunosuppressed individuals over age 35 (or age 45-55 depending on the method) is cost effective at £10 (or £25) per administered dose. Results for younger age-groups, which are only a small proportion of the immunosuppressed population are highly variable, suggesting that uniform vaccination of the entire immunosuppressed population may be the most precautionary option.

Only a few other papers have considered the cost-effectiveness of routine booster vaccination against COVID-19 to-date, although all make quite strong assumptions about population immunity, vaccine protection and waning of protection [25]. A study focusing on the U.S.A., and considering the arrival of new varients, suggested that booster campaigns aimed at the entire population over 5 years of age would be cost effective compared to not boosting [26]. A study in Thailand suggested that universal boosting of the entire population over 18 years of age every two years was more cost-effective than boosting annually, every six months or not at all [27]. A study focused on South Korea compared vaccination of 20% of the entire population with prioritising vaccination of the over 65s, and predicted a switch is the most cost-effective strategy depending on the incidence of infection [28]. Finally, the one paper considering cost-effectiveness of booster vaccination in the UK [29] is part of a series by a team at Moderna [30,31]. This dynamic modelling suggests that vaccination of those over 65 would be highly cost effective using JCVI criteria, even at a price of £74.54 (for vaccine and administration). However, it should be noted that this model assumed high levels of vaccine effectiveness (84% against hospital admission) and very little waning of protection (only declining to 77% after six months). Given this long-duration of protection, we find it surprising that repeated booster vaccines have much appreciable effect.

The level of protection against severe illness that is offered by the vaccine will depend on a number of demographic and epidemiological factors, including: the segregation and mixing patterns of recently-vaccinated and not recently-vaccinated individuals; the criteria applied for admission to hospital; the history of infection and vaccination across the population; and the match between the vaccine under consideration and the circulating sub-variants. All these factors make comparisons between countries and time periods extremely challenging. Throughout, we have considered vaccine effectiveness in terms of the additional relative protection offered by the booster vaccine to the average person receiving it (rather than absolute protection compared to completely unvaccinated individuals), making the history of infection and uptake critical. VIEW-hub [12], provides a comprehensive resource for scientific studies of vaccine effectiveness. In keeping with our analysis, we restrict attention to studies that considered (i) relative protection against hospital admission for COVID-19 in healthy adults (ii) that consider protection over both short and longer time-scales following vaccination (iii) studies from mid-2022 onwards that consider protection against Omicron BA4/5 or later variants; studies that focus on immunosuppressed individuals or that use admission with COVID-like illness have been excluded. For these 28 studies, there is substantial variability in the estimated vaccine effectiveness that could reflect the methodology, the study dates and study population (Supplementary Material, Table S4). Across all studies an early (up to 90 days post vaccination) vaccine effectiveness of 50-60% captures the crude mean, and was within the vast majority of the confidence intervals, whereas later vaccine effectiveness was generally in the range 25-35%. Comparing our estimates to this international picture, we find that early vaccine effectiveness for Estimate 2a was higher (at 66%), later vaccine effectiveness for Estimates 2b and 3a was higher (at 53% and 57%), but Estimate 3b is lower in both periods (at 42% and 15%). These four estimates cover much of the range from the 28 studies, suggesting that our results could have international relevance.

There are some limitations of our analysis, primarily associated with the properties of the data. The partition of the population into just risk groups (no-risk, at-risk but not immunosuppressed and immunosuppressed) is somewhat simplistic. It is likely that a finer partitioning of the risk groups could increase the willingness to pay threshold for some risk groups, although this has to be weighed against the practicalities of more complex criteria. For example, recently diagnosed haematological malignancy or severely reduced kidney function would both place an individual in the at-risk (but not immunosuppressed) group; these conditions have been observed to have a substantially greater impact on the risk of severe outcomes than chronic heart disease which also places an individual in the at-risk group [32,24]. There are also implications if those accepting vaccination are a biased sample of the individual sub-groups (for example, those who perceive themselves at greatest risk may be more likely to be vaccinated). Such biases would reduce the estimated effectiveness from methods 1 and 2. Another potential issue with the data is whether individuals are hospitalised ‘with COVID’ (that is, they have COVID-19 infection at the time of admission, but it is not the cause of admission) or ‘for COVID’ (that is, COVID-19 infection is the primary cause of their admission). Our selection criteria for admissions has been designed to minimise the numbers where COVID-19 is not a major contributing factor; inclusion of individuals ‘with-COVID’ will lead to over-estimates for the willingness to pay threshold.

The static approach we have adopted here provides a worst-case scenario as it does not include the indirect protection offered by vaccination. For example, vaccination of 50-54 year olds only accounts for their direct protection against severe COVID-19 disease, and does not account for the associated reduction in the force of infection experienced by more vulnerable individuals, nor any societal impact due to their illness. However, a dynamic approach where SARS-CoV-2 infection is modelled mechanistically is impractical due to the extreme heterogeneity in history of infection and vaccination. This is exemplified by those studies that have used dynamic SARS-CoV-2 infection transmission models to perform cost-effectiveness analyses of 2023 COVID-19 booster vaccination programmes [26, 28], where heterogeneity is only dependent on vaccine history (with infection history not accounted for).

Our analysis has only considered the cost-effectiveness in terms of severe disease that leads to hospital admission or death. We have not included less severe health burdens, such as illness without medical attendance. Household surveys in England [33] have estimated the prevalence of COVID-19 at around 1% each week during the winter months 2023-2024, peaking at around 2.5-3.0%. However, there is limited data on symptomatic infections and the associated health impact, which is needed to feed into any additional health economic assessment. Therefore, while our willingness to pay represents an underestimate (because less severe health burden is ignored), the scale of this underestimate cannot currently be estimated in the UK.

Furthermore, many unquantifiable uncertainties could influence these findings. We have explored the timing and size of waves (Fig. 4), but cannot associate a probability with any pattern of future outbreaks. These results have shown that while both timing and scale of the epidemic wave(s) have an effect on the vaccine threshold price (with scaling having the largest influence) both are relatively small compared to the impact of age, at most changing eligibility by one 5-year age group for the scale of waves recently experienced. More concerning is the potential impact of novel variants. A novel variant with greater severity or which overcame population-level immunity would likely generate far larger waves of severe cases and therefore merit an expansion of the vaccine programme. Similarly, a new variant with sufficient vaccine escape - such that the level or duration of protection induced by prior COVID-19 vaccination was reduced - may require a re-evaluation of the programme, either reducing its range due to the lower impact per dose, or increasing its range to help combat overall levels of infection and hence protect the most vulnerable.

Vaccination against COVID-19 remains an important public health tool for protecting the elderly and most vulnerable in our society. However, widespread partial immunity in the population, the lower risks of severe illness currently observed, and the need for vaccines to be cost-effective means that offers of population-wide vaccination for all ages are no longer affordable. Instead, we need to carefully balance the vaccine costs against the health saving and health benefits; the precise age threshold will depend on the price paid for the vaccine, but only universal programmes that target those above 70 years old are likely to be cost effective.

## Author contributions

**Matt J. Keeling:** Conceptualisation, Formal analysis, Funding acquisition, Methodology, Software, Validation, Visualisation, Writing - Original Draft, Writing - Review & Editing.

**Edward M. Hill:** Funding acquisition, Methodology, Visualisation, Writing - Original Draft, Writing - Review & Editing.

**Stavros Petrou:** Funding acquisition, Methodology, Writing - Review & Editing.

**Phuong Bich Tran:** Data curation, Formal analysis, Methodology, Writing - Review & Editing.

**May Ee Png:** Data curation, Formal analysis, Methodology, Writing - Review & Editing.

**Sophie Staniszewska:** Funding acquisition, Investigation, Writing - Review & Editing.

**Corinna Clark:** Funding acquisition, Investigation, Writing - Review & Editing.

**Katie Hassel:** Data curation, Investigation, Resources, Writing - Review & Editing.

**Julia Stowe:** Data curation, Investigation, Resources, Writing - Review & Editing.

**Nick Andrews:** Data curation, Investigation, Resources, Writing - Review & Editing.

## Governance

UKHSA has legal permission, provided by Regulation 3 of The Health Service (Control of Patient Information) Regulations 2002, to process patient confidential information for national surveillance of communicable diseases and as such, individual patient consent is not required.

## Ethical approvals

The UK Medicines and Healthcare products Regulatory Agency and the South Central - Berkshire Research Ethics Committee of the Health Research Authority approved the PANORAMIC trial (reference: 21/SC/0393).

## Data Availability

The raw study data are protected and are not available due to data privacy laws. This work is carried out under Regulation 3 of The Health Service (Control of Patient Information) (Secretary of State for Health, 2002) (3) using patient identification information without individual patient consent. Data cannot be made publicly available for ethical and legal reasons, i.e. public availability would compromise patient confidentiality as data tables list single counts of individuals rather than aggregated data.

