## Supporting Information for "Cost-effectiveness of routine COVID-19 adult vaccination programmes in England"

---

#### Table of Contents

**1 Admissions and deaths data by age, risk & vaccination status** **2**

**2 Mathematical outline of the modelling approaches** **3**

**3 Evaluating vaccine programme cost-effectiveness** **5**

**4 International comparison on protection of vaccination against hospitalisation** **8**

**5 Patient and Public Involvement** **9**

---

Here we provide details on the data used throughout this work (Section 1), on the different modelling approaches to infer the health episode outcome rates which are dependent on age, risk and vaccination status (Section 2), and the health economic data and assumption used to make cost-effectiveness assessments (Section 3). When taken together these generate the willingness to pay thresholds for different target groups being included in a routine COVID-19 vaccination programme for England (Section 3). We also place our vaccine effectiveness estimates within a broader international context (Section 4) and overview Patient and Public Involvement in the study (Section 5).

### 1 Admissions and deaths data by age, risk & vaccination status

Our aim was to generate data on the number of individuals each week that were admitted to hospital, admitted to ICU (a term we use for brevity to refer to severe hospitalisation cases) or died with COVID-19. These data are further separated by age (five-year age bins from 15-19 to 90+), risk group (no-risk, at-risk or immunosuppressed, as defined by the Green Book Chapter 14b [1]) and by vaccination status (booster within the last six months or not). This involves the linkage of two data sets on health outcomes (admissions provided by the Secondary Uses Service [2] and deaths provided by the ONS [3]) to individual records on vaccine uptake and demographic status.

The Secondary Uses Service [2] is a database of timely completed hospital admissions, including ICU attendance, for all NHS hospitals in England. To determine COVID-19 related hospitalisations for age 15+ we merged COVID-19 testing data with respiratory coded discharge data obtained from the Secondary User Service. The respiratory discharge ICD-10 codes used were: J04* Acute laryngitis and tracheitis ; J09* Influenza due to identified avian influenza virus; J10* Influenza with pneumonia, other influenza virus identified; J11* Influenza with pneumonia, virus not identified; J12* Viral pneumonia, not elsewhere classified; J13* Pneumonia due to Streptococcus pneumoniae; J14* Pneumonia due to Haemophilus influenzae; J15* Bacterial pneumonia, not elsewhere classified; J16* Pneumonia due to other infectious organisms, not elsewhere classified; J17* Pneumonia in diseases classified elsewhere; J18* Pneumonia, organism unspecified; J20* Acute bronchitis; J21* Acute bronchiolitis; J22* Unspecified acute lower respiratory infection; J80* Adult respiratory distress syndrome ; U071* COVID-19, virus identified; U072* COVID-19, Virus not identified; U04* Severe acute respiratory syndrome (SARS).

We then identified a COVID-19 related hospital admission either by a hospitalisation (with a respiratory coded discharge) that had a COVID-19 positive test (PCR or lateral flow) within the period two days prior to the day after the date of admission, or a discharge where the primary ICD-10 discharge diagnosis was COVID-19.

ICU admissions (corresponding to severe hospitalisation admissions) were those with NHS Office of Population Censuses and Surveys (OPCS) codes for oxygen/ventilation — E85* Ventilation support, E89* Other respiratory support, X58* Artificial support for body system, X52* Oxygen therapy — or Main Specialty/Treatment Function classification associated with an ICU admission (Main Specialty Code: 192 Critical Care Medicine; Treatment Function Code - reporting the specialised service within which the patient was treated: 192 Critical Care Medicine or 242 Paediatric Intensive Care). We collectively refer to these severe hospitalisation admissions as ICU admissions.

We collapsed hospitalisations in the same individuals into 'spells' where there was more than one admission recorded on the same day or where spells overlapped (based on admission and discharge). We combined spells within individuals if two spells started within 15 days of one another. In these combined spells we counted information on severity if it occurred in any of the collapsed admissions.

We identified formal ONS deaths with COVID-19 mentioned on the death certificate [3].

We linked both sets of individuals (admissions and deaths) to UKHSA's national COVID-19 vaccine register, the Immunisation Information System (IIS) [4], that holds COVID-19 vaccine dates, COVID-19 vaccine eligible risk group status [5] and demographic information for all individuals in England. Linkage was performed by matching NHS numbers; matching was perfect for admissions, but failed for around 6% of deaths. The age of these unmatched deaths is known, but vaccination and risk status is randomly assigned based on those with known status.

We derived vaccine coverage data for each week from the IIS using age on 31st August of each year.

#### 2 Mathematical outline of the modelling approaches

We used three differing modelling methodology leading to five different estimates of the protection offered by COVID-19 vaccination. The first method we call the 'aggregate approach', the second method we call the 'time since vaccination approach' and the third method we call the 'literature-based vaccine effectiveness'.

We base each approach on having a set of weekly observations of a particular public health outcome (admission to hospital, admission to ICU or death due to COVID-19), summed across all ages and risk groups; we label this quantity $Y_w$. We also defined the normalised rate of weekly observations:

$$\widehat{Y}_w = \frac{1}{\sum_\omega Y_\omega} Y_w$$

For all that follows we consider a particular age and risk class and a particular observation of a public health outcome; when performing our numerical calculations we need to keep track of these three dimensions, but here for notational brevity we outline how the calculations are performed for a single risk-group, age-class and set of observations. We denote the size of the population as $N$, the number of individuals vaccinated within the last six months as $V_w$ (given the current week being $w$) and the number vaccinated exactly $W$ weeks ago (that is vaccinated in week $w - W$) as $V_w^W$. It should be noted that $\sum_{W<6 \text{ months}} V_w^W = V_w$. For simplicity of notation we define those individuals that have not been vaccinated in the last six months by $U_w = N - V_w$.

For a given observation, we assumed the number observed in a given week ($w$) for a particular public health outcome for those not vaccinated in the last six months was $y_w^U$. Similarly, we assumed the number observed in a given week for a particular public health outcome for those that had been vaccinated in the last six months was $y_w^V$.

##### Method 1: Aggregate approach

Our simplest calculation was to consider the rate $s$ (or $r$) that a random person in the vaccinated (or unvaccinated) group enters our observation class, assuming a Poisson distribution. The probability distribution of a particular rate is proportional to the likelihood:

$$\begin{aligned} P_U(r) &\propto \text{Poisson}\left(\textstyle\sum_w y_w^U | r\overline{U_w}\right) \pi(r) \\ P_V(s) &\propto \text{Poisson}\left(\textstyle\sum_w y_w^V | s\overline{V_w}\right) \pi(s) \end{aligned} \tag{1}$$

with the proportionality constant defined such that the distribution integrates to one. Here $\overline{U_w}$ and $\overline{V_w}$ refer to the average number of unvaccinated and vaccinated individuals during the time period, and $\pi$ is the weakly informative prior probability distribution for the per capita rate of observation ($\pi(r) = exp(-r)$).

Given our assumption that the per capita risks for vaccinated and unvaccinated individuals are independent, the joint distribution is given by the product:

$$\overline{P}_J(r, s) = P_U(r)\, P_V(s) \tag{2}$$

The probability distribution for the vaccine effectiveness $\rho = 1 - s/r$ can then be calculated as:

$$P_{\text{eff}}(\rho) = \int_0^\infty r\overline{P}_J(r, r(1-\rho))dr \tag{3}$$

where the $r$ term in the integral is the natural scaling, and ensures that the probability to integrate to one. This approach effectively treats the entire outbreak as a single observation.

#### Method 2: Time since vaccination approach

Method 2 requires two combined elements to realise the risks and effectiveness distributions: firstly, a method to determine the effectiveness assuming that the level of protection wanes after vaccination; and secondly, a method to include the estimated effectiveness into the rate estimation approach. This two-step process is needed as we combine the estimated effectiveness from older age-groups (which we believe is subject to fewer biases) and use this distribution for all ages.

To acknowledge that the level of protection from the vaccine wanes over time, we define the relative level of protection from the vaccine after $W$ weeks $X^W$, which peaks at one and is assumed to decay substantially by the time when $W$ reaches six months. For Estimate 2a, we assume that $X^W = \exp(-W^2/500)$ which produces a decline similar to that estimated against the initial Omicron BA.1 wave [6]. At any time, we can then define the aggregate level of protection across the entire population $x_w$ in week $w$ as:

$$x_w = \sum_W X^W \frac{V_w^W}{V_w}$$

where $V_w^W$ is the number of individuals vaccinated $W$ weeks ago, and $V_w$ the number vaccinated within the last six months. Given that the level of protection for vaccinated individuals depends on both $s$ (the rate when the vaccine offers maximal protection) and $r$ (the rate when the vaccine offers least protection, assumed to be after six months), it is practical to consider the joint distribution:

$$P_J(r,s) \propto \prod_w \text{Poisson}\left(y_w^U | rU_w\widehat{Y}_w\right) \text{Poisson}\left(y_w^V | (r(1-x_w)+sx_w)V_w\widehat{Y}_w\right) \pi(r)\pi(s) \tag{4}$$

Here, the rate of observation for vaccinated individuals is a weighted combination of $r$ and $s$ depending on the population level amount of protection at any given time, $x_w$. The probability distribution of effectiveness can again be defined from this joint distribution:

$$P_{\text{eff}}(\rho) = \int_0^\infty r P_J(r, r(1-\rho))dr$$

As an extension to this approach, Estimate 2b allowed the decline in protection to be estimated. We define $X^W = \exp(-W^2/(2\sigma^2))$, and set $\sigma$ to maximise Eq. (4) across all age-groups but independently for each public health outcome.

For Method 2 (and hence Estimates 2a and 2b) we combine (by taking the product) the estimated effectiveness, $P_{\text{eff}}(\rho)$, for multiple older age-groups, and once normalised, treat this combined probability ($\pi_{\text{Eff}}(\rho)$) as the effectiveness distribution across all age-groups. The ages chosen to be aggregated, and hence used as a sample for the entire population, correspond to those older age-groups where vaccination was universally offered irrespective of risk status. To calculate the impact of vaccination then requires an approach that can determine the rates consistent with the assumed effectiveness distribution $\pi_{\text{Eff}}(\rho)$.

The joint distribution of the rates $r$ and the effectiveness $\rho$ can then be expressed as:

$$Q_J(r,\rho) = \pi_{\text{Eff}}(\rho)\,\frac{P_J(r, s=r(1-\rho))}{\int P_J(r,s)\,ds} \tag{5}$$

where $P_J$ is defined as in Eq. (4). This allows us to calculate the appropriate joint distribution on the two rates:

$$\widehat{P}_J(r,s) = r\,Q_J(r, 1-s/r) \tag{6}$$

In essence this methodology ensures that the rates for the two groups (vaccinated and unvaccinated) are consistent with both the observations for both classes and the assumed effectiveness distribution.

However, there are younger individuals in the no-risk group who were vaccinated, and therefore do not represent the average population of that age. (There are reasons why these individuals might have been vaccinated including: being pregnant; working in frontline NHS and social care, or care homes; or being at-risk but incorrectly classified in the data). In such cases it is prudent to exclude these individuals from our analysis, in which case:

$$Q_J(r, \rho) = \pi_{\text{Eff}}(\rho)\, P_U(r)$$

where $P_U$ is taken from Eq. (1), and then Eq. (6) still holds.

##### Method 3: Literature-based vaccine effectiveness

For Method 3 (generating Estimates 3a and 3b) the effectiveness has been estimated in advance, which requires a slight adjustment to our previous approach. Usually, the available estimates may be of the effectiveness distribution at different times since vaccination (e.g. $\Pi_{\text{Eff}}(\rho, W)$ in week $W$). Here, in keeping with our previous approaches, we approximate this externally derived distribution with a single distribution ($\pi_{\text{Eff}}(\rho)$) coupled with a deterministic scaling in protection $X^W$ which is also informed by the external data. Using $\pi_{\text{Eff}}$ and $X^W$ based on the external source, we can again return to Eqs. (5) and (6) to inform about the rates.

#### 3 Evaluating vaccine programme cost-effectiveness

For the respective modelling approaches outlined above, the health outcome rates and vaccine effectiveness estimates, we detail the cost-effectiveness assessments. For each of the different target groups being included in a routine COVID-19 vaccination programme for England, this generated a willingness to pay for each administered vaccine (the combined cost of administration and purchase of the vaccine). We first explain the health economic calculations that determined the price point that a vaccine dose would be deemed cost effective. We then overview the use of the health economic parameters within the cost effectiveness assessment.

##### Health economic model calculations

Following from the joint probability density calculations outlined above, we computed the probability density for the health economic benefit ($B = rCost_U - sCost_V$) of a single dose of vaccine as:

$$P_{\text{benefit}}(B) = \int_0^\infty \frac{1}{CostV} P_J\left(r, \frac{rCost_U - B}{CostV}\right) dr \tag{7}$$

For vaccinated and unvaccinated groups respectively, $Cost_V$ and $Cost_U$ represent the total cost associated with whichever severe outcome is being considered (hospital admission, ICU admission or death) and include both direct treatment costs and the value placed on QALY losses. In this formulation, the joint distribution ($P_J$) can be as defined by Eq. (2) (for Method 1) or Eq. (6) (for Methods 2 and 3).

Following JCVI guidelines we considered two quantities [7]. First, using the mean costs and benefits, and assuming one QALY (Quality Adjusted Life Year) is valued at £20,000; we calculated the benefit per vaccine dose at the maximum likelihood values for the rate a random person in the unvaccinated group has the severe outcome under consideration ($r^*$) and the rate a random person in the vaccinated group has the severe outcome ($s^*$):

$$\text{Benefit}_{\text{MLE}} = r^*\, Cost_U \,-\, s^*\, Cost_V \qquad \text{where} \qquad P_J(r^*, s^*) \text{ is the global maximum}$$

Second, using £30,000 per QALY and accounting for variability in cost and rate parameters, we calculated the 10th percentile of the distribution:

$$\text{Benefit}_{10\%} = B \qquad \text{such that} \qquad \int_0^B P_{\text{benefit}}(b)\, db = 0.1$$

A vaccine is then deemed cost effective if the cost per dose including administration is less than both the calculated benefits:

$$\text{Vaccine price} + \text{Admisistration price} \ < \ \min(\text{Benefit}_{\text{MLE}}, \text{Benefit}_{10\%}) \ = \ \text{Willingness to pay threshold.}$$

#### Parameters of the health economic model

The health economic calculations required monetary costs and a measure of disease severity (captured through the loss of Quality-adjusted life years, QALYs) associated with each level of disease outcome. For infection episodes resulting in hospitalisation and ICU admission, we derived estimates from the PANORAMIC trial (Platform Adaptive trial of NOvel antiviRals for eArly treatMent of COVID-19 in the Community) [8] (Table S3).

*PANORAMIC trial.* The purpose of the PANORAMIC trial was to find out in which people new antiviral treatments for COVID-19 in the community could reduce the need for hospital admission and improve health outcomes. Participants enrolled in the PANORAMIC trial had tested positive for SARS-CoV-2 infection, had ongoing symptoms consistent with COVID-19, had not been previously hospitalised due to COVID-19 (a criteria resulting from the trial focus being on community-based treatment) and were either aged 50 years or older, or were aged between 18 to 49 years old and considered clinically vulnerable. Although some of these criteria may produce a bias in terms of rates of hospital admission, given we only use to PANORAMIC trial to estimate the cost of hospital episodes any bias should be minimal.

We further limited the eligible participants for the calculation of our health economic parameter estimated. This step comprised limiting to participants from England (85.3% of the trial population), participants who had received at least one dose of vaccination, and participants who were treated with either molnupiravir or usual care.

We grouped participants into the following health states based on the reporting of any pre-existing medical conditions: Immunocompromised - participant-reported having a weakened immune system; Not immunocompromised but high risk - participant-reported lung disease, heart disease, diabetes, or obesity; Not at risk - participant reported not having any of the aforementioned medical conditions.

*Cost calculations: Hospital and ICU admissions.* In our analysis we assumed the raw hospitalisation (or ICU costs) for vaccinated and unvaccinated individuals to be equal, with only the associated length of stay ($LoS_U$ and $LoS_V$) varying.

Hospital admission costs were a function of the age of the individual and their risk status:

$$Cost_X(a, r, \text{hospital}) \ = \ C_h(a, r) \times LoS_X(a, r) \ + \ QALY \times Q_h(a, r)$$

where $X \in \{U, V\}$ and $QALY$ is the monetary value associated with one QALY loss (either £20,000 for our central estimate or £30,000 when considering uncertainty). We had age and risk dependent per day cost, $C_h$, and loss of QALYS, $Q_h$, estimated from the PANORAMIC study (Fig. S1, left-hand column).

For costs associated with ICU admission we made a similar calculation:

$$Cost_X(a, r, \text{ICU}) \;=\; C_I(a, r) \times LoS_X(a, r) \;+\; QALY \times Q_I(a, r)$$

with per day cost, $C_I$, and loss of QALYS, $Q_I$, again being age and risk dependent (Fig. S1, right-hand column).

We took hospital admission unit costs from the NHS 2022/23 National Cost Collection Data Publication [9], then attached the appropriate Healthcare Resource Group (HRG) code to each patient to get their hospitalisation (or ICU) cost per episode. The cost of each patient was then divided by the number of days in the hospital (or ICU) to get the hospitalisation (or ICU) cost per day. We computed costs over six months, adjusted by trial allocation group and sex. We analysed adjusted values using a generalised linear model with log-link.

We valued QALYs using EQ-5D-5L and UK utility values, derived using the approach recommended by NICE [10]. This approach applies a validated mapping function onto the UK EQ-5D-3L tariff set that has been developed by the NICE Decision Support Unit [10]. We adjusted QALYs lost by trial allocation group, sex and baseline EQ-5D-3L utility score. We analysed adjusted QALYs using beta-regression.

For hospital and ICU admissions we factored in the uncertainty in the monetary cost and QALY-loss estimates. The uncertainty in QALY losses was especially substantial for those admitted to ICU (Fig. S1).

**Cost calculations: Deaths.** For infection episodes resulting in mortality (Fig. S1, lower panel), we assumed no monetary costs and assumed that QALY losses ($Q_D$) depended on both age and risk group:

$$Cost_X(a, \text{death}) \;= QALY \times Q_D(a, r)$$

The quality-adjusted life-year losses associated with COVID-19 deaths ($Q_D(a, r)$) were sourced from the literature. These accounted for QALY losses at various ages and adjusted for the presence of comorbidities that influence both life-expectancy and health-related quality of life [11]. The standard life expectancy estimation approach, which focuses on conditional life expectancy upon reaching a specific age, were modified to incorporate: (i) the impact of comorbidities on life expectancy using the standardized mortality ratio ($SMR$); (ii) the effect of pre-existing comorbidities on health-related quality of life ($qCM$) to estimate QALYs over time; and (iii) discounting ($\varepsilon = 0.035$ throughout) [11].

The following parameters were assumed for different risk groups: for the immunocompromised group, defined as participants reporting a weakened immune system ($SMR = 2$; $qCM = 0.8$); for the not immunocompromised but high risk group—e.g., those reporting lung disease, heart disease, diabetes, or obesity ($SMR = 1.5$; $qCM = 0.9$); and for the low-risk group—participants without the mentioned medical conditions ($SMR = 1$; $qCM = 1$). Since these are arbitrary thresholds, we reviewed relevant literature to check their appropriateness and found that our assumptions generally aligned with established findings [12–14]. However, it is important to note that multimorbidity and immunosuppression exist on a spectrum concerning their effects on mortality and morbidity. Specifically, individuals with severe conditions like stroke or multiple chronic diseases may exhibit higher $SMR$ and lower $qCM$ values. Similarly, immunocompromised individuals often have additional comorbidities, suggesting that a lower $qCM$ threshold could also be considered for this group. However, making slight adjustments to these parameters is unlikely to significantly alter the model outcomes.

The final QALY losses associated with mortality for each age and risk group are shown in Fig. S1 (lower panel).

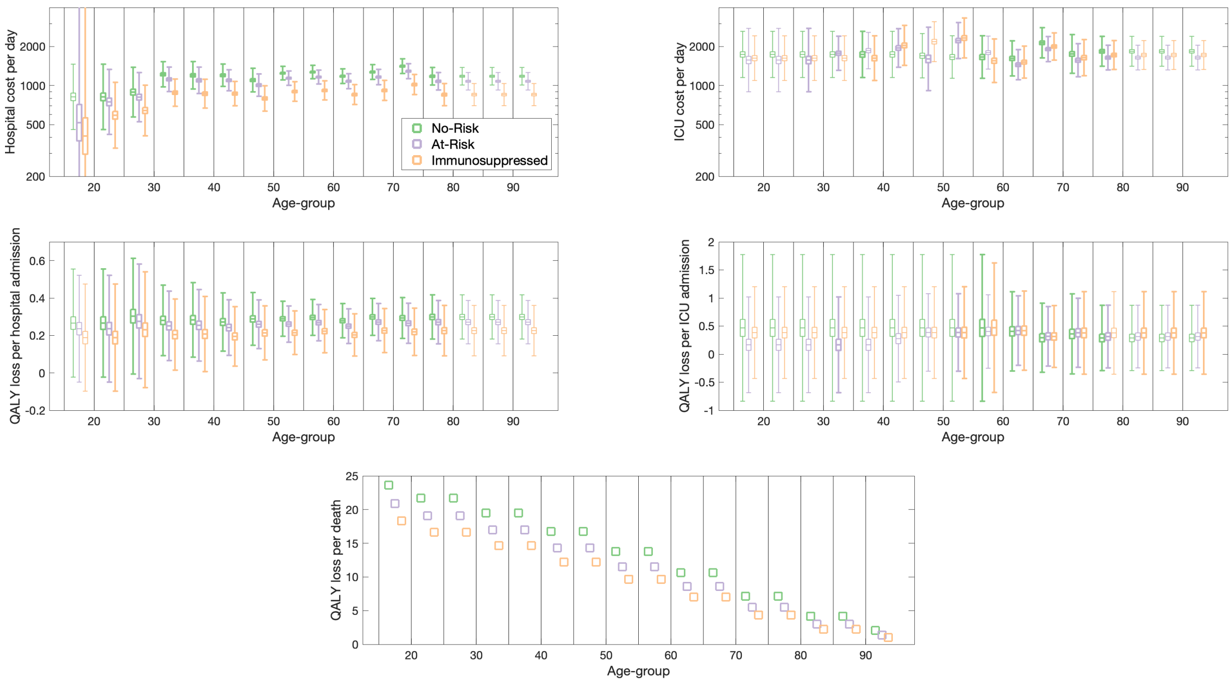


**Fig. S1: Health episode costs and QALY losses (together with uncertainties) used in the health economic assessment.** All data is from the PANORAMIC (Platform Adaptive trial of NOvel antiviRals for eArly treatMent of COVID-19 in the Community) trial [8]dataset of 2484 individuals with symptoms consistent with COVID-19. We estimated the healthcare costs per day from the raw data using a generalised linear model (GLM) with log-link. We estimated the hospitalisation and ICU QALY losses using beta regression. We estimated the QALY loss from premature death using risk-group specific increases in mortality and lower quality of life estimates [11] and a discounting of 3.5% per year. We colour code all the estimated values by risk group status: no-risk (green, left of each triplet group); at-risk but not immunosuppressed (purple, centre of each triplet group); and immunosuppressed (orange, right of each triplet group); fainter box-whisker plots are for age/risk-groups where there were no data and displayed results are extrapolations.

### 4 International comparison on protection of vaccination against hospitalisation

It is important to place the estimates used within the main paper (Figure 2) within a broader international context. In this regard, we use VIEW-hub [15], a collaboration between the International Vaccine Access Center, W.H.O. and the Coalition for Epidemic Preparedness Innovations, which provides a comprehensive resource for scientific studies of vaccine effectiveness. We note that the estimated variance depends on multiple epidemiological and demographic elements. Firstly, the baseline to which the additional booster dose is compared; while it might be simpler to compare newly vaccinated to completed unvaccinated individuals, throughout our work it is more intuitive to compare recently vaccinated to not-recently vaccinated. Secondly, the effectiveness of the vaccine is likely to depend on the history of infection and vaccination which will differ between individuals and between regions and times. Finally, effectiveness will depend on the match between the vaccine and the dominant variants, which again will likely depend on region and times. In keeping with our analysis, we restrict attention to studies that consider (i) relative protection against hospital admission for COVID-19 in healthy adults (ii) that consider protection over both short and longer time-scales following vaccination (iii) later studies from mid-2022 onwards that consider protection of booster doses against Omicron BA4/5 or later variants. Studies that focus on immunosuppressed individuals, that use admission with COVID-like illness have been excluded, or that were conducted early in the outbreak are excluded. This leaves 28 studies, mainly from the USA and Europe (Table S4).

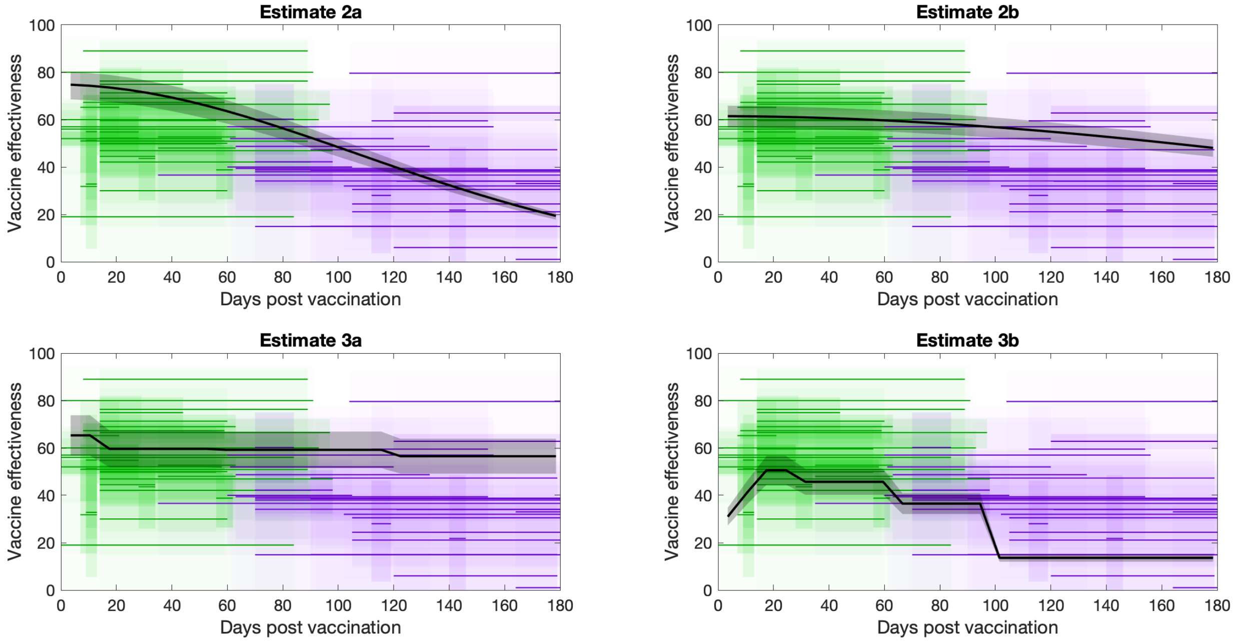


**Fig. S2: Comparison between the estimates of vaccine effectiveness against hospitalisation (for recently vaccinated versus not recently vaccinated) used in the paper and estimates from 28 international studies**. We took data from the VIEW-hub [15] and focused on protection in healthy adults against hospital admission against Omicron variants from mid-2022 onwards, where there are multiple estimates of vaccine effectiveness at different time-scales post vaccination. Early vaccine effectiveness, shortly after vaccination, is coloured green, later vaccine effectiveness is coloured purple, estimates used in the paper are in black; confidence intervals are shaded to illustrate the certainty at each point.

In Fig. S2 we compare the four estimates used in this paper (in black) to early (green) and late (purple) from the 28 studies identified in VIEW-hub [15]. We use shading to highlight the uncertainty from each estimate at each time and value. Compared to these international estimates our four estimates show some discrepancies. Estimate 2a is slightly higher than the international average in the first 90 days; Estimates 2b and 3a are higher than the average post 90 days; whereas Estimate 3b may be lower than the average throughout. Potentially, these differences reflect different populations, different vaccination and infection histories, and different SARS-CoV-2 variants. What is reassuring is that the estimates used in the main paper often bracket the international estimates, providing additional confidence that our results have international relevance.

### 5 Patient and Public Involvement

As part of the MEMVIE programme of research, which seeks to embed patient and public voices within vaccination modelling studies [16], we presented the models from this study to a standing group of six public contributors. We presented the model methodology and findings in an accessible narrative form by the modelling team, allowing the Patient and Public Involvement (PPI) group to question the assumptions, parameters, and findings. Due to time constraints on the modelling team, in this instance, the discussion took place after (rather than during) model development; however, the PPI discussions form part of ongoing work to further refine the MEMVIE Framework for Public Involvement in Mathematical and Economic Modelling, and to facilitate PPI implementation within vaccination modelling.

### Supplementary Tables

**Table S1: Willingness to pay thresholds for the Spring 2023 booster**. Thresholds are given for all ages (although ages 15-39 have been amalgamated for simplicity), for no-risk, at-risk and universal vaccine deployment and for the five estimates of vaccine effectiveness. Values are rounded to the nearest pound, and values less than £1 are indicated. Ages, risk-groups and estimates where the willingness to pay threshold is determined by the JCVI uncertainty condition (that there is a 90% chance of cost-effectiveness at £30,000 per QALY) are marked with an asterisk. Shaded cells correspond with ages and risk groups where vaccination was recommended in the Spring 2023 COVID-19 vaccination programme [17].

| | Universal | | | | | No Risk | | | | | At Risk | | | | |
|---|---|---|---|---|---|---|---|---|---|---|---|---|---|---|---|
| Age | E1 | E2a | E2b | E3a | E3b | E1 | E2a | E2b | E3a | E3b | E1 | E2a | E2b | E3a | E3b |
| **15-39** | - | < £1 ⋆ | < £1 ⋆ | £1 | < £1 | - | < £1 ⋆ | < £1 ⋆ | < £1 | < £1 | < £1 | £1 ⋆ | £1 ⋆ | $\leq$ £2 | £1 |
| **40-44** | - | < £1 | < £1 ⋆ | £1 | < £1 | - | < £1 | < £1 ⋆ | < £1 | < £1 | < £1 | £1 | £1 | £3 | £1 |
| **45-49** | - | £1 | < £1 | £1 | £1 | - | < £1 ⋆ | < £1 ⋆ | £1 | < £1 ⋆ | < £1 | £2 | £1 | £5 | £2 |
| **50-54** | - | £1 | £1 | £2 | £1 | - | < £1 | < £1 | £1 | < £1 | < £1 | £3 | £2 | £7 | £4 |
| **55-59** | - | £2 | £1 | £3 | £2 | - | £1 | £1 | £1 | < £1 | £1 ⋆ | £4 | £3 | £8 | £4 |
| **60-64** | - | £2 | £1 | £5 | £2 | - | £1 | £1 | £2 | £1 | < £1 | £4 | £3 | £10 | £5 |
| **65-69** | £1 ⋆ | £6 | £4 | £11 | £6 | < £1 | £2 | £1 | £3 | £2 | £2 ⋆ | £10 | £7 | £20 | £11 |
| **70-74** | £3 ⋆ | £7 | £5 | £16 | £8 | £3 ⋆ | £3 | £2 | £5 | £3 | £3 ⋆ | £11 | £7 | £26 | £13 |
| **75-79** | £27 | £16 | £11 | £34 | £17 | £21 | £8 | £7 | £13 | £7 | £32 | £20 | £13 | £48 | £23 |
| **80-84** | £33 | £20 | £13 | £50 | £24 | £25 | £11 | £10 | £20 | £9 | £36 | £24 | £15 | £62 | £30 |
| **85-89** | £36 | £38 | £26 | £92 | £44 | £15 ⋆ | £27 | £24 ⋆ | £48 | £24 | £39 | £41 | £26 | £103 | £50 |
| **90+** | £19 ⋆ | £36 | £22 | £101 | £47 | < £1 | £31 | £27 | £53 | £27 | £35 | £37 | £21 | £111 | £51 |

**Table S2: Willingness to pay thresholds for the Autumn 2023 boosters**. Thresholds are given for all ages, for no-risk, at-risk and universal vaccine deployment and for the five estimates of vaccine effectiveness. Values are rounded to the nearest pound, and values less than £1 are indicated. Ages, risk-groups and estimates where the willingness to pay threshold is determined by the JCVI uncertainty condition (that there is a 90% chance of cost-effectiveness at £30,000 per QALY) are marked with an asterisk. Shaded cells correspond with ages and risk groups where vaccination was recommended in the Autumn 2023 COVID-19 vaccination programme [18].

| | Universal | | | | | No Risk | | | | | At Risk | | | | |
|---|---|---|---|---|---|---|---|---|---|---|---|---|---|---|---|
| Age | E1 | E2a | E2b | E3a | E3b | E1 | E2a | E2b | E3a | E3b | E1 | E2a | E2b | E3a | E3b |
| **15-19** | - | < £1 | < £1 | < £1 | < £1 | - | < £1 | < £1 | < £1 | < £1 | < £1 | £2 * | £2 * | £3 | £2 * |
| **20-24** | - | < £1 | < £1 * | < £1 | < £1 | - | < £1 | < £1 * | < £1 | < £1 | < £1 | £1 * | £1 * | £2 * | £1 * |
| **25-29** | - | < £1 * | < £1 * | £1 * | < £1 * | - | < £1 * | < £1 * | < £1 * | < £1 * | < £1 | £1 * | £1 * | £2 * | £1 * |
| **30-34** | - | £1 | < £1 | £1 | < £1 | - | < £1 | < £1 | < £1 | < £1 | < £1 | £2 | £1 | £3 | £1 |
| **35-39** | - | £1 * | £1 * | £1 * | £1 * | - | < £1 * | < £1 * | < £1 * | < £1 * | < £1 | £2 | £2 | £4 | £2 |
| **40-44** | - | £1 | £1 | £1 | £1 | - | < £1 | < £1 | < £1 | < £1 | < £1 | £5 | £4 | £7 | £4 |
| **45-49** | - | £2 | £1 | £2 | £1 | - | £1 | £1 | £1 | £1 | < £1 | £5 | £4 | £7 | £3 |
| **50-54** | - | £3 | £3 | £4 | £2 | - | £1 | £1 | £1 | £1 | < £1 | £9 | £8 | £13 | £7 |
| **55-59** | - | £5 | £4 | £6 | £3 | - | £2 | £2 | £2 | £1 | < £1 | £12 | £11 | £17 | £9 |
| **60-64** | - | £7 | £6 | £10 | £5 | - | £3 | £2 | £3 | £2 | £5 * | £15 | £13 | £22 | £11 |
| **65-69** | £14 | £16 | £14 | £21 | £10 | £7 | £6 | £6 | £6 | £3 | £24 | £29 | £25 | £41 | £19 |
| **70-74** | £29 | £24 | £22 | £34 | £15 | £11 | £10 | £10 | £10 | £4 | £45 | £37 | £33 | £57 | £25 |
| **75-79** | £74 | £51 | £48 | £80 | £30 | £41 | £25 | £27 | £25 | £10 | £95 | £67 | £62 | £114 | £43 |
| **80-84** | £84 | £72 | £68 | £128 | £45 | £49 | £42 | £45 | £43 | £16 | £98 | £83 | £77 | £162 | £57 |
| **85-89** | £169 | £134 | £125 | £228 | £83 | £119 | £89 | £95 | £94 | £36 | £182 | £146 | £133 | £263 | £95 |
| **90+** | £148 | £141 | £132 | £275 | £97 | £19 * | £97 | £99 | £100 | £45 | £172 | £150 | £138 | £309 | £107 |

**Table S3: Summary of health economic values for hospitalisation, ICU and death events.** We provide the following statistics stratified by age-group and risk status, which are displayed in Fig. S1. For infection episodes resulting in hospitalisation and ICU admission, we report the mean estimates (with 95% confidence intervals in parentheses) for the cost per day (£) to the nearest integer and QALY loss to 2 decimal places. Estimates in regular typeface are from the PANORAMIC trial (Platform Adaptive trial of NOvel antiviRals for eArly treatMent of COVID-19 in the Community) [8], with estimates in italic typeface corresponding to age and risk-groups where there were no data and displayed results are extrapolations. Per death we report the QALY loss to 2 decimal places, with estimates produced using the approach of Briggs *et al.* [11] to adjust for the presence of comorbidities that influence both life-expectancy and health-related quality of life.

| | | **Hospitalisation** | | **ICU** | | **Death** |
|---|---|---|---|---|---|---|
| **Age** | **Risk status** | **Cost per day (£)** | **QALY loss** | **Cost per day (£)** | **QALY loss** | **QALY loss** |
| **15-19** | **Not at risk** | *819 (459,1463)* | *0.27 (-0.02,0.56)* | *1739 (1155,2620)* | *0.47 (-0.84,1.77)* | 23.61 |
| | **At-risk** | 518 (32,8331) | *0.24 (-0.05,0.52)* | *1574 (897,2762)* | *0.17 (-0.69,1.02)* | 20.89 |
| | **Immunosuppressed** | 409 (26,6573) | *0.19 (-0.10,0.47)* | *1629 (1095,2421)* | *0.38 (-0.43,1.20)* | 18.31 |
| **20-24** | **Not at risk** | 819 (459,1463) | 0.27 (-0.02,0.56) | *1739 (1155,2620)* | *0.47 (-0.84,1.77)* | 21.72 |
| | **At-risk** | 749 (421,1334) | 0.24 (-0.05,0.52) | *1574 (897,2762)* | *0.17 (-0.69,1.02)* | 19.08 |
| | **Immunosuppressed** | 591 (330,1060) | 0.19 (-0.10,0.47) | *1629 (1095,2421)* | *0.38 (-0.43,1.20)* | 16.62 |
| **25-29** | **Not at risk** | 891 (573,1386) | 0.30 (-0.01,0.61) | *1739 (1155,2620)* | *0.47 (-0.84,1.77)* | 21.72 |
| | **At-risk** | 815 (527,1261) | 0.28 (-0.03,0.58) | 1574 (897,2762) | *0.17 (-0.69,1.02)* | 19.08 |
| | **Immunosuppressed** | 643 (411,1008) | 0.23 (-0.08,0.54) | *1629 (1095,2421)* | *0.38 (-0.43,1.20)* | 16.62 |
| **30-34** | **Not at risk** | 1223 (978,1530) | 0.28 (0.09,0.47) | *1739 (1155,2620)* | *0.47 (-0.84,1.77)* | 19.52 |
| | **At-risk** | 1119 (900,1390) | 0.25 (0.07,0.44) | 1776 (1312,2405) | 0.17 (-0.69,1.02) | 16.96 |
| | **Immunosuppressed** | 883 (693,1124) | 0.20 (0.01,0.39) | 1629 (1095,2421) | *0.38 (-0.43,1.20)* | 14.64 |
| **35-39** | **Not at risk** | 1202 (942,1533) | 0.28 (0.08,0.48) | 1739 (1155,2620) | *0.47 (-0.84,1.77)* | 19.52 |
| | **At-risk** | 1099 (869,1389) | 0.25 (0.06,0.45) | *1862 (1348,2571)* | *0.17 (-0.69,1.02)* | 16.96 |
| | **Immunosuppressed** | 867 (671,1120) | 0.21 (0.01,0.41) | 1628 (1095,2421) | *0.38 (-0.43,1.20)* | 14.64 |
| **40-44** | **Not at risk** | 1203 (987,1467) | 0.27 (0.12,0.43) | *1739 (1155,2620)* | *0.47 (-0.84,1.77)* | 16.73 |
| | **At-risk** | 1100 (913,1325) | 0.24 (0.09,0.39) | 1951 (1385,2748) | *0.17 (-0.69,1.02)* | 14.30 |
| | **Immunosuppressed** | 868 (700,1077) | 0.20 (0.04,0.35) | 2043 (1433,2912) | *0.38 (-0.43,1.20)* | 12.19 |
| **45-49** | **Not at risk** | 1104 (895,1361) | 0.29 (0.15,0.43) | *1739 (1155,2620)* | *0.47 (-0.84,1.77)* | 16.73 |
| | **At-risk** | 1009 (828,1230) | 0.26 (0.13,0.39) | 1607 (916,2819) | *0.28 (-0.49,1.05)* | 14.30 |
| | **Immunosuppressed** | 797 (636,998) | 0.21 (0.07,0.36) | *2181 (1530,3109)* | *0.38 (-0.43,1.20)* | 12.19 |
| **50-54** | **Not at risk** | 1249 (1107,1409) | 0.29 (0.20,0.38) | *1700 (1147,2520)* | *0.47 (-0.84,1.77)* | 13.79 |
| | **At-risk** | 1142 (1005,1297) | 0.26 (0.16,0.36) | 2225 (1617,3062) | 0.39 (-0.30,1.08) | 11.52 |
| | **Immunosuppressed** | 901 (758,1072) | 0.21 (0.10,0.33) | 2330 (1635,3319) | 0.38 (-0.43,1.20) | 9.64 |
| **55-59** | **Not at risk** | 1273 (1131,1434) | 0.30 (0.20,0.39) | 1661 (1139,2424) | 0.47 (-0.84,1.77) | 13.79 |
| | **At-risk** | 1164 (1029,1317) | 0.27 (0.17,0.37) | *1797 (1340,2409)* | *0.40 (-0.25,1.06)* | 11.52 |
| | **Immunosuppressed** | 919 (776,1089) | 0.22 (0.11,0.34) | 1556 (1057,2289) | 0.47 (-0.68,1.63) | 9.64 |
| **60-64** | **Not at risk** | 1183 (1039,1347) | 0.28 (0.19,0.37) | 1621 (1187,2214) | 0.41 (-0.30,1.11) | 10.64 |
| | **At-risk** | 1082 (947,1236) | 0.25 (0.16,0.34) | 1450 (1110,1895) | 0.42 (-0.20,1.04) | 8.61 |
| | **Immunosuppressed** | 854 (716,1019) | 0.20 (0.09,0.32) | 1518 (1143,2017) | 0.42 (-0.29,1.12) | 7.04 |
| **65-69** | **Not at risk** | 1274 (1117,1452) | 0.30 (0.20,0.40) | 2139 (1635,2797) | 0.29 (-0.32,0.91) | 10.64 |
| | **At-risk** | 1165 (1017,1334) | 0.27 (0.17,0.37) | 1913 (1531,2390) | 0.32 (-0.21,0.85) | 8.61 |
| | **Immunosuppressed** | 919 (769,1098) | 0.22 (0.11,0.34) | 2003 (1574,2548) | 0.32 (-0.24,0.87) | 7.04 |
| **70-74** | **Not at risk** | 1413 (1240,1609) | 0.29 (0.18,0.40) | 1755 (1245,2474) | 0.36 (-0.35,1.07) | 7.14 |
| | **At-risk** | 1292 (1131,1476) | 0.27 (0.16,0.37) | 1570 (1170,2106) | 0.38 (-0.24,1.00) | 5.50 |
| | **Immunosuppressed** | 1020 (853,1219) | 0.22 (0.09,0.35) | 1643 (1192,2265) | 0.38 (-0.36,1.11) | 4.34 |
| **75-79** | **Not at risk** | 1182 (1011,1381) | 0.30 (0.18,0.42) | 1839 (1411,2397) | 0.29 (-0.29,0.87) | 7.14 |
| | **At-risk** | 1080 (925,1262) | 0.27 (0.16,0.39) | 1645 (1321,2048) | 0.31 (-0.24,0.87) | 5.50 |
| | **Immunosuppressed** | 853 (701,1038) | 0.23 (0.09,0.36) | 1722 (1332,2226) | *0.38 (-0.36,1.11)* | 4.34 |
| **80-84** | **Not at risk** | *1182 (1011,1381)* | *0.30 (0.18,0.42)* | *1839 (1411,2397)* | *0.29 (-0.29,0.87)* | 4.15 |
| | **At-risk** | *1080 (925,1262)* | *0.27 (0.16,0.39)* | *1645 (1321,2048)* | *0.31 (-0.24,0.87)* | *3.00* |
| | **Immunosuppressed** | *853 (701,1038)* | *0.23 (0.09,0.36)* | *1722 (1332,2226)* | *0.38 (-0.36,1.11)* | 2.25 |
| **85-89** | **Not at risk** | *1182 (1011,1381)* | *0.30 (0.18,0.42)* | *1839 (1411,2397)* | *0.29 (-0.29,0.87)* | 4.15 |
| | **At-risk** | *1080 (925,1262)* | *0.27 (0.16,0.39)* | *1645 (1321,2048)* | *0.31 (-0.24,0.87)* | 3.00 |
| | **Immunosuppressed** | *853 (701,1038)* | *0.23 (0.09,0.36)* | *1722 (1332,2226)* | *0.38 (-0.36,1.11)* | 2.25 |
| **90+** | **Not at risk** | *1182 (1011,1381)* | *0.30 (0.18,0.42)* | *1839 (1411,2397)* | *0.29 (-0.29,0.87)* | 2.07 |
| | **At-risk** | *1080 (925,1262)* | *0.27 (0.16,0.39)* | *1645 (1321,2048)* | *0.31 (-0.24,0.87)* | 1.39 |
| | **Immunosuppressed** | *853 (701,1038)* | *0.23 (0.09,0.36)* | *1722 (1332,2226)* | *0.38 (-0.36,1.11)* | 1.00 |

**Table S4: Estimates of vaccine effectiveness (VE) against hospital admission from 28 studies** [15]. Papers were chosen that included at least two estimates of VE at different periods post vaccination, that focused on later booster doses, and that compared those recently vaccinated to those that have not received the booster dose. Results are also given for the four estimates in the paper that include declining VE.

| Short Term | | Long Term | | | |
|---|---|---|---|---|---|
| VE | Days | VE | Days | Ages | Reference |
| 52% (49-55%) | 0-58 | 32% (5-52%) | 102+ | >18 | [19] |
| 57% (48-64%) | 0-92 | 21% (-15-46%) | 184-270 | >50 | [20] |
| 60% (52-67%) | 0-93 | 28% (-5-50%) | 184-271 | >65 | [20] |
| 44.5% (35.4-52.2%) | 14-34 | 24.4% (10.3-36.3%) | 105+ | >75 | [21] |
| 69% (50-82%) | 14-29 | 40% (19-55%) | 60-105 | >18 | [22] |
| 50.9% (45.1-56.1%) | 14-89 | 47.3% (32-59.1%) | 90-179 | 65-79 | [23] |
| 42.1% (36.4-47.2%) | 14-90 | 38.6% (17.4-54.3%) | 90-180 | >80 | [23] |
| 52.2% (41.3-61.1%) | 14-28 | 59.5% (21.4-79.1%) | 112-154 | >18 | [24] |
| 59.6% (47.2-69%) | 14-56 | 48.7% (-0.7-72.5%) | 63-133 | >18 | [25] |
| 76.3% (21.4-92.9%) | 14-89 | -1.2% (-73.9-41.1%) | 270-364 | >18 | [26] |
| 65.2% (50.6-79.6%) | 7-21 | 60.2% (45.3-75%) | 70-84 | >65 | [27] |
| 54.8% (46.8-61.6%) | 14-28 | 42.2% (32.3-50.6%) | 70-98 | >65 | [28] |
| 80% (50-94%) | 14-89 | 15% (-12-35%) | 90-179 | >60 | [29] |
| 50% (15-71%) | 14-59 | 57% (30-73%) | 60-156 | >18 | [30] |
| 71.3% (44.9-85.1%) | 14-60 | 52% (-1.2-77.3%) | 61-120 | >6 months | [31] |
| 19% (-8-39%) | 0-84 | 1% (-53-36%) | 164+ | >80 | [32] |
| 56% (37-69%) | 0-84 | 33% (10-50%) | 164+ | 65-79 | [32] |
| 43.6% (26-57.1%) | 28-34 | 28% (3.6-46.2%) | 112-119 | >12 | [33] |
| 38.8% (26.4-49.2%) | 56-62 | 21.8% (0-48.6%) | 140-146 | >12 | [33] |
| 80% (15-95%) | 0-91 | 38% (-37-71%) | 92-212 | >18 | [34] |
| 54.9% (21.7-74%) | 9-13 | 38.8% (13.8-56.6%) | 70+ | >75 | [28] |
| 51.2% (26.3-67.7%) | 9-13 | 14.9% (-8.3-33.1%) | 70+ | >75 | [28] |
| 66.5% (59.8-72.2%) | 14-97 | 33.8% (7.1-52.9%) | 182+ | 65-79 | [35] |
| 46.9% (29.3-53.6%) | 14-98 | 29.8% (11.3-44.4%) | 182+ | >80 | [35] |
| 69% (62-74%) | 14-63 | -5% (-77-37%) | 238-287 | >75 | [36] |
| 52.7% (24.6-70.4%) | 35-63 | 21.1% (9.6-31.1%) | 105+ | >50 | [37] |
| 48% (38.5-56%) | 35-63 | 30.5% (18.7-40.6%) | 105+ | >50 | [37] |
| 89% (51-66%) | 8-89 | 31% (11-44%) | 180+ | >60 | [38] |
| 74.9% (66.4-81.3%) | 14-44 | 79.6% (43.2-92.7%) | 104+ | >6 | [39] |
| 51% (45-57%) | 7-59 | 6% (-7-18%) | 120-179 | >18 | [40] |
| 49.8% (24.8-61%) | 14-56 | 39.4% (21.7-52.7%) | 63-154 | >12 | [41] |
| 31.8% (15.5-44.9%) | 7-13 | 38% (31-44.3%) | 70+ | >50 | [42] |
| 51% (37.6-61.5%) | 7-13 | 34.1% (29.2-38.7%) | 70+ | >50 | [42] |
| 50% (38-60%) | 14-60 | 34% (-383-91%) | 121-180 | >65 | [43] |
| 30% (-48-67%) | 14-60 | 1% (-55-37%) | 241-300 | >65 | [43] |
| 32.8% (5.5-52.3%) | 9-13 | 36.6% (14.7-52.9%) | 35+ | >75 | [44] |
| 67.3% (63.9-70.4%) | 8-59 | 62.8% (59.3-65.9%) | 120+ | >18 | [45] |
| 66.4% (60.9-71.2%) | 0-90 | 34.2% (31.4-36.7%) | 91-180 | >65 | Estimate2a |
| 60.1% (55.4-64.4%) | 0-90 | 53% (48.8-56.8%) | 91-180 | >65 | Estimate2b |
| 60.3% (52.5-68.2%) | 0-90 | 57.3% (49.9-64.8%) | 91-180 | >65 | Estimate3a |
| 42.1% (37.1-47.2%) | 0-90 | 15.4% (13.5-17.2%) | 91-180 | >65 | Estimate3b |

[31] Ackerson BK, Bruxvoort KJ, Qian L, Sy LS, Qiu S, *et al.* Effectiveness and durability of mRNA-1273 BA. 4/BA. 5 bivalent vaccine (mRNA-1273.222) against SARS-CoV-2 BA. 4/BA. 5 and

XBB sublineages. *Human Vaccines & Immunotherapeutics* **20**(1):2335052 (2024).
[32] Fontán-Vela M, Kissling E, Nicolay N, Braeye T, Van Evercooren I, *et al.* Relative vaccine effectiveness against COVID-19 hospitalisation in persons aged ≥65 years: results from a VEBIS network, Europe, October 2021 to July 2023. *Eurosurveillance* **29**(1):2300670 (2024).
[33] Paritala S, Xu Y, Du Y, Donahue M, Maloney P, *et al.* Effectiveness of Bivalent Boosters Over Nine and Half Months. *Journal of Biotechnology and Biomedicine* **6**(4):585–589 (2023).
[34] Tartof SY, Slezak JM, Puzniak L, Hong V, Frankland TB, *et al.* Effectiveness of BNT162b2 BA. 4/5 bivalent mRNA vaccine against a range of COVID-19 outcomes in a large health system in the USA: A test-negative case–control study. *The Lancet Respiratory Medicine* **11**(12):1089–1100 (2023).
[35] Machado A, Kislaya I, Soares P, Magalhaes S, Rodrigues AP, *et al.* Bivalent mRNA vaccine effectiveness against COVID-19 infections, hospitalisations and deaths in Portugal: a cohort study based on electronic health records, September 2022 to May 2023. *medRxiv* pages 2023–09 (2023).
[36] Stecher M, Kristoffersen AB, Lie K, Andersen SR, Meijerink H, *et al.* Effectiveness and durability of a second COVID-19 booster against severe outcomes among older people in Norway: a population-based cohort study comparing mono-and bivalent booster doses. *International Journal of Epidemiology* **52**(6):1716–1724 (2023).
[37] Kirsebom FCM, Harman K, Lunt RJ, Andrews N, Groves N, *et al.* Vaccine effectiveness against hospitalisation estimated using a test-negative case-control study design, and comparative odds of hospital admission and severe outcomes with COVID-19 sub-lineages BQ. 1, CH. 1.1. and XBB. 1.5 in England. *The Lancet Regional Health–Europe* **35** (2023). doi:10.1016/j.lanepe.2023.100755.
[38] Wee LE, Pang D, Chiew C, Tan J, Lee V, *et al.* Long-term real-world protection afforded by third mRNA doses against symptomatic severe acute respiratory syndrome coronavirus 2 infections, coronavirus disease 19-related emergency attendances and hospitalizations amongst older Singaporeans during an Omicron XBB wave. *Clinical Infectious Diseases* **77**(8):1111–1119 (2023).
[39] Tseng HF, Ackerson BK, Sy LS, Tubert JE, Luo Y, *et al.* mRNA-1273 bivalent (original and Omicron) COVID-19 vaccine effectiveness against COVID-19 outcomes in the United States. *Nature Communications* **14**(1):5851 (2023).
[40] Link-Gelles R. Estimates of bivalent mRNA vaccine durability in preventing COVID-19–associated hospitalization and critical illness among adults with and without immunocompromising conditions—VISION Network, September 2022–April 2023. *MMWR. Morbidity and Mortality Weekly Report* **72** (2023).
[41] Lin DY, Xu Y, Gu Y, Zeng D, Sunny SK, *et al.* Durability of bivalent boosters against Omicron subvariants. *New England Journal of Medicine* **388**(19):1818–1820 (2023).
[42] Kirsebom FCM, Andrews N, Stowe J, Ramsay M, Bernal JL. Duration of protection of ancestral-strain monovalent vaccines and effectiveness of bivalent BA. 1 boosters against COVID-19 hospitalisation in England: a test-negative case-control study. *The Lancet Infectious Diseases* **23**(11):1235–1243 (2023). doi:10.1016/S1473-3099(23)00365-1.
[43] Poukka E, Perälä J, Nohynek H, Goebeler S, Auranen K, *et al.* Relative effectiveness of bivalent boosters against severe COVID-19 outcomes among people aged ≥65 years in Finland, September 2022 to August 2023. *Eurosurveillance* **29**(37):2300587 (2024).
[44] UKHSA. COVID-19 vaccine surveillance report, July 2024 (2024). URL https://assets.publishing.service.gov.uk/media/669923b20808eaf43b50d1fd/Vaccine_surveillance_report_2024_week_29.pdf.
[45] Ranzani OT, Hitchings MD, de Melo RL, de França GV, Fernandes CdFR, *et al.* Effectiveness of an inactivated Covid-19 vaccine with homologous and heterologous boosters against Omicron in Brazil. *Nature communications* **13**(1):5536 (2022).
